# Optimized ITS/CITS models for intervention evaluation considering the nonlinear impact of covariates

**DOI:** 10.1101/2023.03.27.23287776

**Authors:** Xiangliang Zhang, Rong Yin, Yan Pan, Wenfang Zhong, Di Kong, Wen Chen

**Author notes:** Corresponding author: Wen Chen, PhD. First co-author (Xiangliang Zhang, Rong Yin).

## Abstract

There is a lack of approaches to evaluate the effectiveness of interventions when there are nonlinear impacts of covariates to the outcome series.

Based on the classic framework of ITS/CITS segmented regression, while considering autocorrelation of time series, we adopted a nonlinear dynamic modeling strategy (Hammerstein) to measure the nonlinear effects of covariates, and proposed four optimized models: ITS-A, CITS-A, ITS-HA, and CITS-HA. To compare the accuracy and precision in estimating the long-term impact of an intervention between the optimized and classic segmented models, we constructed a sequence generator to simulate the outcome series with actual characteristics. The relative error with respect to the true value was the accuracy indicator, and the width of the 95% CI and the truth value coverage rate of the corresponding 95% CI are the precision indicator for model assessments.

The relative error of impact evaluation in the four optimized models was 4.49 percentage points lower than that in the classic models, specifically ITS-A (14.34%) and ITS-HA (21.47%) relative to ITS (26.66%), CITS-A (16.57%), and CITS-HA (17.94%) relative to CITS (21.59%). The width of the 95% CI of point estimate of long-term impacts in the optimized models was 0.1261, which was expanded by 58.71% compared with 0.0875 for the classic model. However, the optimized models covered the true value in all test scenarios, whereas the coverage rates of the classic ITS and CITS models were 73.33% and 83.33%, respectively.

The optimized models are useful tools as they can assess the long-term impact of interventions with additional considerations for the nonlinear effects of covariates and allow for modeling of time-series autocorrelation and lag of intervention effects.

## 1. Introduction

An interrupted time series design[1] is a major approach to the evaluation of interventions when randomized controlled trials (RCTs) are not feasible. [2–4] According to the presence or absence of a control group in the study, the interrupted time series design can be divided into two categories: interrupted time series without a control group (ITS) and interrupted time series with a control group (controlled interrupted time series, CITS). Compared with ITS, CITS is more capable of eliminating bias caused by unknown factors that are often accompanied by interventions, and thus provides stronger evidence for causal inferences.[1]

Segmented regression,[5] which fits a linear regression model based on time-series data, is commonly applied in ITS/CITS design. It reduces bias and precisely assesses the effects of an intervention by controlling for covariates. Although widely used, it is based on strict assumptions, such as linearity, independence, normality, and homogeneity of variance.[6] In segmented regression analysis, the effects of an intervention are usually considered to be direct, immediate, and leapfrogged, which is not consistent with reality;[7, 8] only the linear effects of covariates can be considered.[9]

However, in some cases, time-series data do not satisfy all the assumptions above. First, some covariates in ITS/CITS have a nonlinear effect on the outcome series.[10–12] For example, previous studies have found nonlinear effects of socioeconomic factors on infectious and non-communicable diseases, as well as on health care utilization.[13–15] Second, when an intervention has gradual or delayed impacts on the outcome series, a lag along with its lagged pattern of effects, rather than direct effects, should be considered. For example, lag periods have been reported in dozens of drug utilization studies.^17^ The effects of the Medicare Act on hospital admission tended to emerge gradually after its implementation.^16^ Moreover, substantial evidence has proven the hysteresis effects of air pollution exposure on the mortality and morbidity of respiratory and cardiovascular diseases.[16–19] Meanwhile, different patterns of lagged intervention effects exist,[20] particularly in epidemiological research.[21] Third, time series are inherently autocorrelated, in which case the regression, forecast, and further evaluation would be inaccurate using segmented regression analysis.[22–24]

In the situations mentioned above, classic segmented regression fails to address such methodological issues because of the challenges of specifying a model. To tackle the problem of autocorrelation, the autoregressive integrated moving average (ARIMA) model is feasible; however, it does not consider the effects of covariates.[25] Furthermore, ARIMAX, an extended ARIMA model, seems more applicable because it controls for the linear effects of covariates by introducing exogenous variables *X*. In other words, by incorporating ARIMAX into the analytical frameworks of ITS/CITS, we can obtain a mixed model, denoted as ITS-ARIMAX (ITS-A)/CITS-ARIMAX(CITS-A), by which we can deal with the situation when both autocorrelation and linear effects of covariates occur.[26] In addition, lagged effects in ITS-A/CITS-A models can be tackled by employing different ramp functions, such as the ReLU function (linear function) and sigmoid function.[27]

However, there is still a lack of sufficient solutions for the nonlinear effects of covariates. To address this, a nonlinear dynamic modeling strategy using the Hammerstein model would be helpful. The Hammerstein model, which is a typical modular nonlinear model, consists of 1) a sub-nonlinear system with memoryless dynamics, and 2) a linear sub-system with memory in series.[28] In some fields, the linear module of the Hammerstein model has been set as an ARX or ARIMA structure to solve nonlinear dynamic modeling problems with serial autocorrelation.[29, 30] There have been studies in economics that modeled dependencies between the volatility risk indexes of different countries; however, there have been no attempts at intervention evaluation yet.^31^ Therefore, the combination of the Hammerstein model and the ITS-A/CITS-A model, which is denoted as ITS-Hammerstein-ARIMAX (ITS-HA) and CITS-Hammerstein-ARIMAX (CITS-HA), respectively, would hopefully help to measure the nonlinear effects of covariates.

In this study, we simulated data with the following characteristics: nonlinear effects of covariate series, different lag patterns of intervention effects, and autocorrelation of outcome series. Based on the data, we estimated the accuracy and precision of the six models mentioned above (ITS-segmented regression, CITS-segmented regression, ITS-A, CITS-A, ITS-HA, and CITS-HA) for evaluation of intervention effects, to verify whether the optimized models (ITS-A, CITS-A, ITS-HA, and CITS-HA) are superior to the classic models (ITS-segmented regression, and CITS-segmented regression), as well as their applicability.

## 2. Models

### 2.1 Classic segmented Regression models

#### 2.1.1 Interrupted Time Series (ITS) - segmented regression

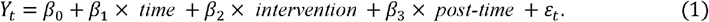

In the above model, *Y*_*t*_ is the value of the outcome series at time point *t. time* is an indicator variable of the time point, ranging from the first observation point to the last observation point (*time* = 0, 1, 2, …, *n* − 1). *intervention* was a dummy variable for intervention implementation. Here, 0 and 1 represent the values before and after the intervention, respectively. *post-time* was an indicator variable for recording the time since the intervention was implemented. The value of *post-time* before intervention is set as 0 and increases over time during the post-intervention period (*post-time* = 0, 1, 2, …, *m, m* < *n* − 1). *ε*_*t*_ is the random error term at time *t. β*_1_ represents the baseline trend of the outcome series before the intervention. *β*_2_ reflects the instantaneous impact of the intervention on *Y*_t_. *β*_3_ is the change in the trend (slopes) of the outcome series due to the intervention and reflects the long-term impact of the intervention.

#### 2.1.2 Controlled Interrupted Time Series (CITS) - segmented regression

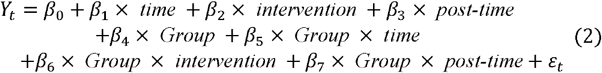

In the above model, *Group* is a grouping dummy variable that takes 0 for the control group and 1 for the intervention group. The other variables have the same meaning as in Equation 1. *β*_4_ is an intercept term indicating the difference between the control and intervention groups before intervention. *β*_5_ indicates the distinction between the trends (slopes) of the two groups before intervention. *β*_6_ reflects the difference in the instantaneous changes in *Y*_*t*_ between the two groups after the intervention. *β*_7_ is the difference between the trends (slopes) of the two groups after the intervention, reflecting the long-term impact of the intervention.

### 2.2 Optimized models

Based on classic segmented regression models (mentioned in 2.1), we proposed a general approach (Equation 3) to optimize intervention evaluation by considering the nonlinear effects of covariates, lag of intervention effects, and autocorrelation of outcome series. 1) We used the structure of the Hammerstein model by combining a nonlinear module with linear modules represented by ITS-A/CITS-A to control for the nonlinear effects of covariates. 2) We incorporated the lag of intervention effects into the model by introducing a ramp function f(t). 3) The introduction of the ARIMA term can address the autocorrelation of the time series itself. Then, we provide a general expression of the optimized model as follows:

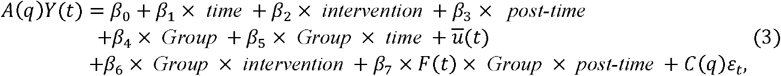

where

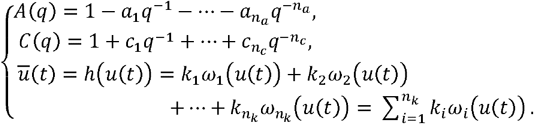

Here, *A*(*q*) and *C*(*q*) are the components of the ARIMA model. *q* is the time lag operator, that is, *q*^−*i*^ *Y*(*t*) = *Y*(*t* − *i*). *ū*(*t*) represents the component of the Hammerstein model, and *h*(*u*(*t*)) is the specific form of the corresponding nonlinear module. In this study, we chose a polynomial function (Equation 4), which is commonly used as the specific form of *h*(*u*(*t*)), namely

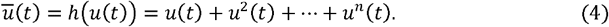

where *n* is the nonlinear order of the covariate effect *u*(*t*) is the corresponding covariate sequence. In addition, *F*(*t*) in Equation 3 is a piecewise function (Equation 5) that is embedded to fit the specific lag pattern of the intervention effects.

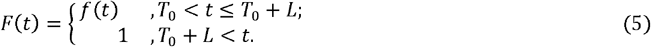

Here, *T*_0_ is the starting point for the intervention. *L* is the assumed maximum lag. For the ramp function *f*(*t*), there are plenty of optional forms, for example, sigmoid, hyperbolic tangent, and ReLU, Mish.[27] The selection of its form depends on the actual lag pattern of intervention effects. In this study, we chose three commonly used patterns:1) *ReLu*: *f*(*t*) =*t* denoted by LagF_ReLu, 2) 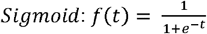 denoted by LagF_Sig, and 3) no lag effects: *f*(*t*) = 1 denoted by LagF_Null.

In the framework of Equation 3, we construct different models when considering the linear or nonlinear effects of the covariates. Specifically, we presented the following optimized models:1) **ITS-A** (Equation 6): The ITS-A model takes into account both autocorrelation of outcome series *Y*(*t*) and linear effects of covariate sequence *u*(*t*) without the control group; 2) **ITS-HA** (Equation 7): The ITS-HA model considers both autocorrelation of outcome series *Y*(*t*) and nonlinear effects of covariate sequence *u*(*t*); 3) **CITS-A** (Equation 8): The CITS-A model includes the control group on the basis of ITS-A; and 4) **CITS-HA** (Equation 9): The CITS-A models include the control group based on ITS-HA.

#### ITS- ARIMAX (ITS-A)

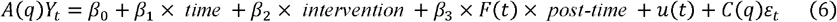

#### ITS-Hammerstein-ARIMAX (ITS-HA)

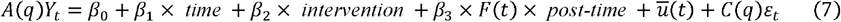

#### CITS- ARIMAX (CITS-A)

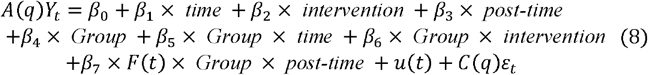

#### CITS- Hammerstein-ARIMAX (CITS-HA)

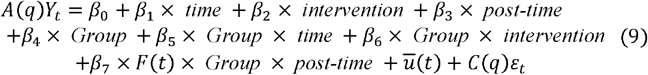

Assignment rules of variables (*time, intervention, post-time, and Group*) and indicating meanings of coefficients in the optimized models (Equations 6-9) are identical to those in the ITS/CITS model (Equations 1-2). In other words, *β*_2_ indicates instant impacts, *β*_3_ indicates long-term impacts in ITS-A and ITS-HA (ITS model class), *β* indicates instant impacts, and *β* indicates long-term impacts in CITS-A and CITS-HA (CITS model class). In this study, we only focused on the accuracy and precision of the optimized models in assessing the long-term impacts of the intervention, that is, *β*_3_ for the ITS model class and *β*_7_ for the CITS model class.

### 2.3 Data Simulation

To generate simulated data that satisfy the aforementioned characteristics, including the nonlinear effects of covariates, different lagged patterns of intervention effects, and autocorrelation of outcome series, we constructed a sequence generator.

We first set the length of the simulated series with 60-time points (both covariate sequence and outcome series) and divided the series equally into two parts, representing the pre-intervention and post-intervention phases. Considering the total length of the post-intervention time series, we set the maximum length of the lagged effect of the intervention to seven time points, approximately one-fourth the length of the post-intervention time series.

In our simulations, the outcome series *Y*_*t*_ was generated by a nonlinear transformation of covariate sequence *u(t)* (Detail in 2.3.1 *Data simulation for outcome series - Nonlinear effects of the covariate*). We compared the differences between the pre- and post-intervention phases of the covariate sequence to simulate the intervention effect of the outcome series. In the study, the pre- and post-intervention series for covariate generation were defined as *p*(*t*) = 0.01*t* + 1 and *q*(*t*) = 0.02*t* + 1, respectively. In addition, we added a white noise series *N*(0,0.1) to *p*(*t*) and *q*(*t*) to simulate random perturbation of covariate sequence *u*(*t*), and then obtained covariate sequences *u*(*t*)*c*_1_, *u*(*t*)*c*_2_ for control groups and *u*(*t*)*Int, u*(*t*)*Relu, u*(*t*)*Sig* for intervention groups (**Fig 1** *State* ①).

**Figure 1.**
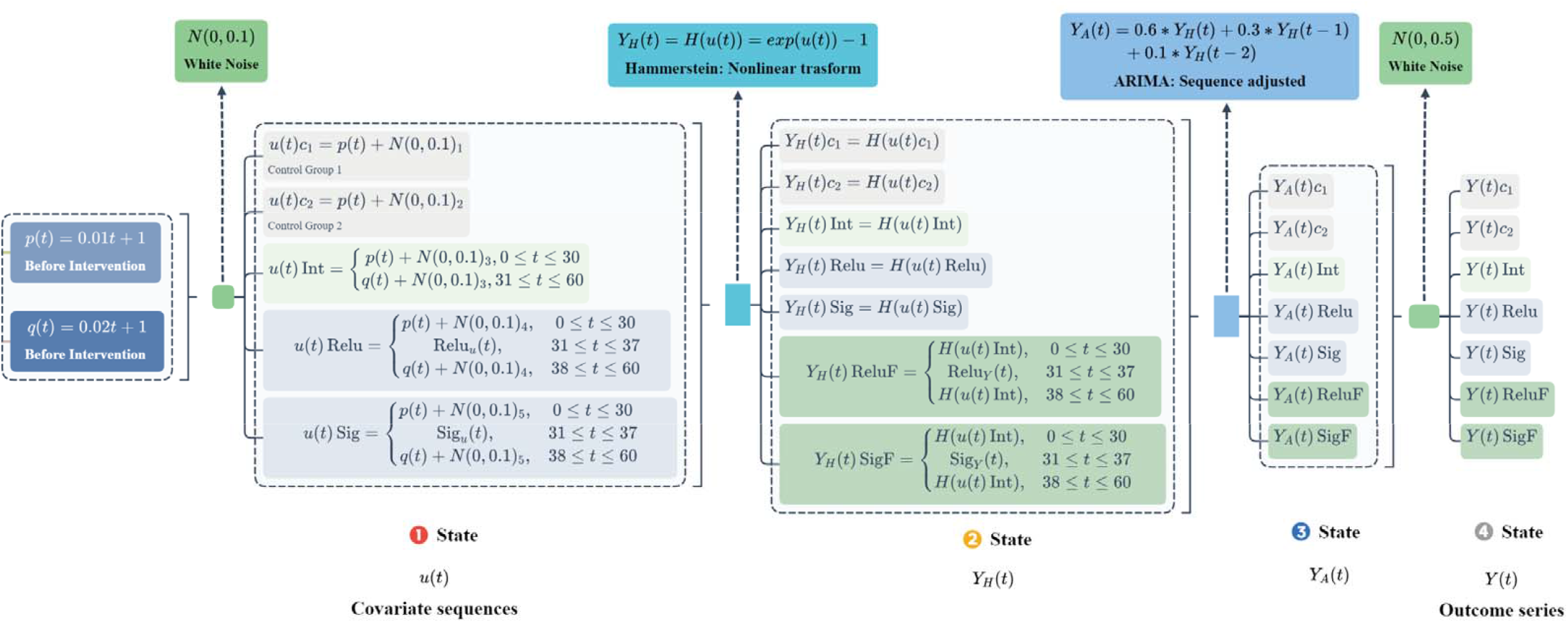
Outcome series generation flowchart.

#### 2.3.1 Data simulation for outcome series

The data simulation process of the outcome series is introduced in **Fig 1**.

##### Nonlinear effects of the covariate

To achieve a nonlinear effect of the covariates on outcome series *Y*(*t*), we performed a nonlinear transformation of the covariate sequence *u*(*t*), that is, *Y*_*H*_(*t*) = *H*(*u*(*t*)) = *exp*(*u*(*t*))− 1, into *Y*_*H*_(*t*) in *State* ② (**Fig 1**).

##### Different lag patterns of intervention effects

In this study, we considered three typical lagged patterns: no lag, linear *Relu* lag, and nonlinear sigmoid lag. When there was no lag after the intervention, the covariate sequence was denoted as *u*(*t*)*Int* and the outcome series was denoted as *Y*(*t*)*Int*. The lag of the intervention effect could be stimulated by a lag term of covariate sequence *u*(*t*) or by a lag term of *Y*_*H*_(*t*), which means that we could simulate the lag of the intervention effect by interpolating the covariate sequence either *Y*(*t*) or *Y*_*H*_(*t*) after the intervention point. Different interpolation modes can simulate lagged patterns. We used linear and S-smooth interpolation to simulate *Relu* and Sigmoid lagged patterns separately. These two interpolations of covariate sequence *u*(*t*) were expressed by *Relu*_*u*_(*t*) (Equation 10) and *Sig*_*u*_(*t*) (Equation 11) (**Fig 1** *State* ①).

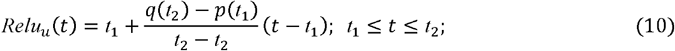

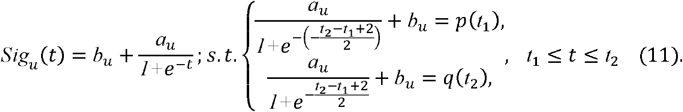

Correspondingly, the two interpolations of *Y*_*H*_(*t*) are expressed by *Relu*_*Y*_(*t*) (Equation 12) and *Sig*_*Y*_(*t*) (Equation 13) (**Fig 1** *State* ②).

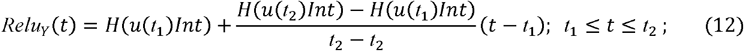

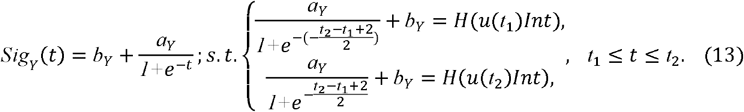

Subsequently, *Y*_*H*_(*t*) *Relu*_*Y*_, *Y*_*H*_(*t*)*Sig* (by lag terms of *u*(*t*)) and *Y*_*H*_(*t*) *ReluF, Y*_*H*_(*t*) *SigF* (by lag terms of *Y*_*H*_(*t*)) were generated (**Fig 1** *State* ②).

##### Autocorrelation

To simulate the autocorrelation of the outcome variable *Y* (*t*), we adjusted *Y*_*H*_(*t*) using the equation *Y*_*A*_(*t*) = 0.6 * *Y*_*H*_(*t*) + 0.3 * *Y*_*H*_(*t*−1) + 0.1 * *Y*_*H*_(*t*−2), so that *Y*_*A*_(*t*) (**Fig 1** *State* ③) was autocorrelated.

We then added a white noise series *N* (0,0.5) to *Y*_*A*_(*t*) to simulate random perturbation of outcome series *Y* (*t*), and obtained the final outcome series: *Y*_*A*_(*t*)*Int, Y* (*t*)*Relu, Y* (*t*)*Sig, Y* (*t*) *ReluF, Y* (*t*)*SigF*.

Along with the simulation process, we generated two control groups (*Y* (*t*)*c*_1_ and *Y* (*t*)*c*_2_), without simulating the intervention effects. The series of two control groups were identical, except for the random numbers of simulation noise. All data generated by the sequence generator are shown in **Supplementary materials Fig S1**.

### 2.4 Model processing - coefficient identification

#### 2.4.1 Illustration on the covariate included in the model calculation

In the simulation process, different outcome series were generated separately, based on different covariate sequences. For the intervention group, the differences between the pre- and post-intervention phases of the covariate sequence formed the intervention effect. However, when evaluating the impact of the intervention effect using the six models, we must assume that the covariate sequence maintains its trend and maintains the same nonlinear relationship with the outcome series. Otherwise, it was impossible to judge whether the change in outcome series after the intervention was due to the intervention or the covariate. Therefore, for covariate sequences of the intervention group, which were incorporated into the model for calculation, we selected the pre-intervention phase of the covariate sequence (the first 30 data points of *u*(*t*)*Int, u*(*t*)*Relu, u*(*t*)*Sig*) and mean post-intervention phase of covariate sequence (the last 30 data points of *u*(*t*)*c*_1_ and *u*(*t*)*c*_2_) to form complete covariate sequences for the final model calculation.

#### 2.4.2 Software

MATLAB (version 9.6.0.10727) and Tableau Desktop 20021.3 were used in our numerical simulations and calculations, as well as in the graphical drawing of the results.

### 2.5 Model evaluation

In this study, models were judged based on their accuracy and precision in estimating the long-term impacts of intervention (*β*_3_ in ITS models and *β*_7_ in CITS models). We calculated the true values of long-term impacts on five different outcome series (*Y Int, Y Relu, Y Sig, Y ReluF, Y SigF*). The relative error with respect to the true value was the accuracy indicator, and the width of the 95% CI and the truth value coverage rate of the corresponding 95% CI was the precision indicator for model assessments.

#### 2.5.1 True value of long-term impacts

Referring to the generation logic of the simulated data in Section 2.3 (without considering the white noise of different outcome series), we employed a least squares method to estimate the slope values in the regression models: *β*_*pre*_ (before intervention) and *β*_*post*_ (after intervention). We subtracted the pre-intervention slope *β*_*pre*_ from the post-intervention slope *β*_*post*_ to obtain the true value of the long-term impact of the intervention, that is, *β*_*true*_ = *β*_*post*_ − *β*_*pre*_. **Table 1** shows the true value of the long-term impact (the last column) for different outcome series, with a detailed calculation of *β*_*pre*_ and *β*_*post*_ for different outcome series in **Supplementary materials Explanation M1**.

**Table 1.** The true value of long-term impact for different outcome series.

| | $\beta_{pre}$ | $\beta_{post}$ | $\beta_{postL}$ | $\beta_{postU}$ | $F$ | $P$ | $Residual$ | $\beta_{true}$ |
| --- | --- | --- | --- | --- | --- | --- | --- | --- |
| **Y(t)Int** | 0.0318 | 0.1363 | 0.1322 | 0.1404 | 4699.7 | <0.001 | 0.0089 | 0.1045 |
| **Y(t)Relu** | 0.0318 | 0.1641 | 0.1562 | 0.1719 | 1828.8 | <0.001 | 0.0331 | 0.1323 |
| **Y(t)Sig** | 0.0318 | 0.1643 | 0.1531 | 0.1755 | 905.5 | <0.001 | 0.0670 | 0.1325 |
| **Y(t)ReluF** | 0.0318 | 0.1608 | 0.1537 | 0.1679 | 2151.8 | <0.001 | 0.0270 | 0.1290 |
| **Y(t)SigF** | 0.0318 | 0.1621 | 0.1512 | 0.1730 | 929.2 | <0.001 | 0.0636 | 0.1303 |

Note: $\beta_{post} L$ and $\beta_{post} U$ are the lower and upper limits, respectively, of the 95% CI for the $\beta_{post}$ estimate. $F$ , $P$ , and $Residual$ are model statistics. The $F$ is the test statistic of the F-test in the regression model, and $P$ is the corresponding p-value. $Residual$ is an estimate of error variance.

#### 2.5.2 Possible scenarios for evaluation

The possible scenarios for evaluating the different models were distinct (**Fig 2**). For ITS, we considered all possible scenarios under the condition of five different outcome series above *Y(t)* and three different ramp functions *f*(*t*), with 3 × 5 = 15 scenarios in total. For the ITS-A, we additionally considered the autocorrelation of the outcome series. However, according to the generation process of the sequence, we only considered its second-order autocorrelation, that is, *Y*(*t*) had a relationship with *Y*(*t* − 1) and *Y*(*t* − 2) as the model embedding of ARIMA. Thus, the number of possible scenarios for ITS-A was 15. For the ITS-HA, the nonlinearity of the covariates was additionally considered in the model. In our study, we chose polynomial functions (Equation 4) as the nonlinear structure of the covariate sequence *u*(*t*) and tested different polynomial function orders, *n* (*n* = 2 − 10). Thus, the number of possible scenarios for ITS-HA was 135 (3 × 5 × 9 = 135) under the condition of five different outcome series *Y*(*t*), three different forms of the ramp function *f*(*t*) and nine different nonlinear orders *n* in *u*(*t*).

**Fig 2.**
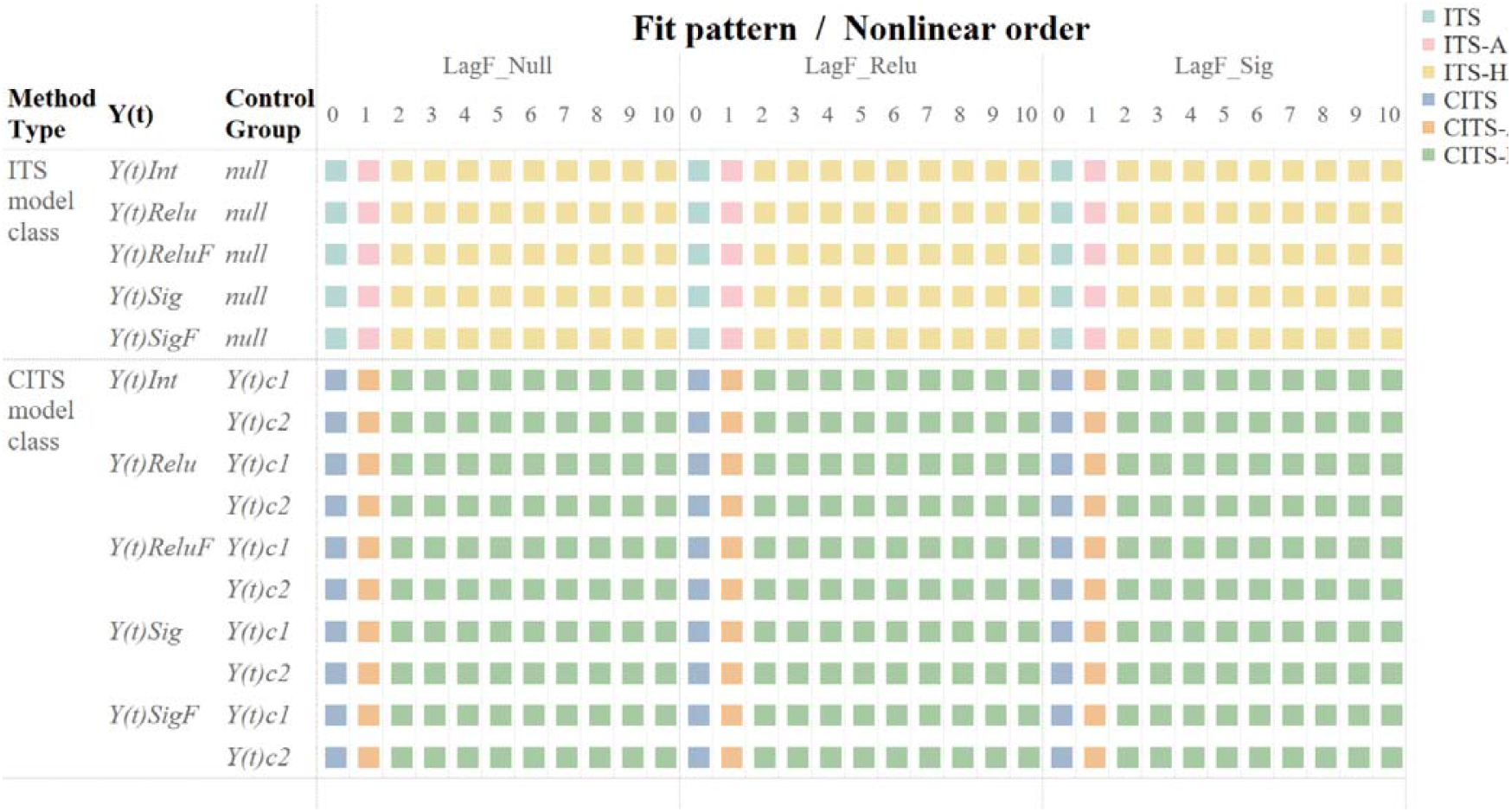
Possible scenarios for evaluation of different models. Note: The six different colors represent the six different models. Each box represents a specific evaluation scenario.

Additionally, for CITS, CITS-A, and CITS-HA, we evaluated the intervention effect with two different control groups (*Y*(*t*)*c*_1_ and (*Y*(*t*)*c*_2_). Therefore, the number of possible scenarios doubled compared to ITS, ITS-A, and CITS-HA, namely, 3 ×5 ×2 = 30 for CITS, CITS-A, and 3 ×5 ×9 ×2 = 270 for CITS-HA. Overall, the total number of scenarios considered in this study was 495.

#### 2.5.3 Accuracy and precision indicator

We used the relative error 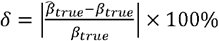 as the accuracy indicator. 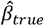 is the point estimate value of the long-term impacts in a model (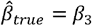 in the ITS model class and 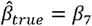 in the CITS model class). Absolute errors 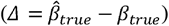 could be used to compare the relative magnitudes of the model point estimates to the true values. Meanwhile, the width of the 95% CI of 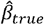 and the actual coverage rate of the CIs served as precision indicators in this study.

For a certain model, the mean relative error and mean width of the CI are the averages of the relative error and interval widths weighted by various possible scenarios.

## 3. Results

### 3.1 Accuracy of evaluation

#### 3.1.1 Optimized models VS. Classic segmented regression models

We compared the accuracy of the evaluation of the long-term impacts of an intervention for each model based on the five outcome series, *Y*(*t*). As shown in **Table 2**, the mean relative errors varied considerably across the models. The optimized models showed higher accuracy than the classic segmented regression models. Specifically, for the ITS model class, both ITS-A (mean relative error:14.34%) and ITS-HA (mean relative error:21.47%) showed a lower mean relative error than ITS-segmented regression (mean relative error:26.66%). Similarly, for the CITS models, both CITS-A (16.57%) and CITS-HA (17.94%) exhibited a lower mean relative error than the corresponding CITS-segmented model (21.59%).

**Table 2.** The relative error for different models.

| Model | Control Group | LagF_Null (%) | LagF_Relu (%) | LagF_Sig (%) | Mean relative error (%) |
| --- | --- | --- | --- | --- | --- |
| ITS | - | 29.68 | 24.77 | 25.53 | 26.66 |
| ITS-A | - | 16.33 | 13.12 | 13.57 | 14.34 |
| ITS-HA | - | 26.34 | 18.89 | 19.17 | 21.47 |
| CITS | $Y(t)c_1$ | 5.13 | 4.68 | 4.15 | 4.65 |
| | $Y(t)c_2$ | 42.22 | 36.20 | 37.16 | 38.53 |
|  | Mean | 23.68 | 20.44 | 20.66 | 21.59 |
| CITS-A | $Y(t)c_1$ | 7.91 | 9.94 | 9.05 | 8.97 |
| | $Y(t)c_2$ | 28.68 | 21.50 | 22.35 | 24.18 |
|  | Mean | 18.30 | 15.72 | 15.70 | 16.57 |
| CITS-HA | $Y(t)c_1$ | 8.61 | 9.90 | 8.80 | 9.10 |
| | $Y(t)c_2$ | 30.89 | 24.32 | 25.16 | 26.79 |
|  | Mean | 19.75 | 17.11 | 16.98 | 17.94 |

Note: “-” means no data because there were no control groups for the ITS model class (ITS, ITS-A, and ITS-HA). LagF\_Null (%), LagF\_Relu (%), and LagF\_Sig (%) show the percentage of relative error when fitting three lag patterns: no lag, linear lag, and nonlinear lag of the intervention effects, respectively.

Meanwhile, the point estimates of long-term impacts in most scenarios tended to be larger than their true values, as their absolute errors were mostly greater than zero (**Fig 3** and **Fig S2-S6**).

**Figure 2.**
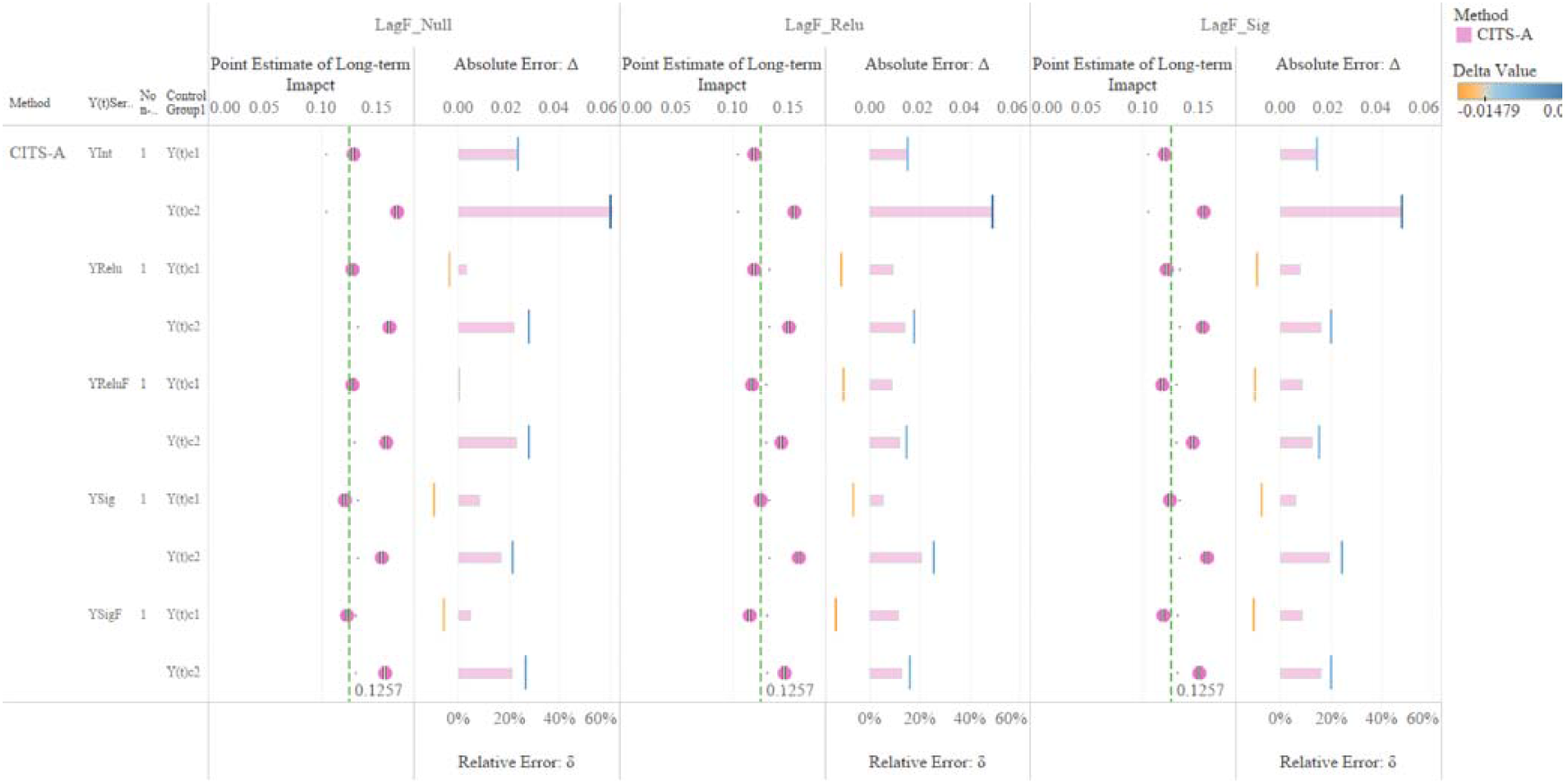
Point estimates of long-term impact based on the CITS-A model and their corresponding error rate. The vertical columns represent three lag patterns: no lag (LagF_Null), linear lag (LagF_Relu), and nonlinear lag (LagF_Sig). Each column is divided into two sub-vertical columns. In the left sub-vertical column, the dots represent the point estimate of the long-term impacts of an evaluation based on CITS-A. The green, thin, and dashed lines represent the mean of the true values of the long-term impacts. In the right sub-vertical column, the height of each bar represents the magnitude of the relative error δ, and the broken vertical lines represent the absolute error Δ.

#### 3.1.2 CITS model class VS. ITS model class

In general, the CITS model classes (CITS, CITS-A, and CITS-HA) performed better in accurately evaluating the long-term impacts of an intervention than the ITS model classes (ITS, ITS-A, and ITS-HA), and the improvement in accuracy was particularly noticeable when *Y*(*t*)*c*_1_ was chosen as the control group. For the CITS model class, the mean relative error was 18.15%, which was lower than that for the ITS model class (21.29%).

#### 3.1.3 CITS model class with *Y*(*t*)*c*_1_ VS. CITS model class with *Y*(*t*)*c*_2_

For the CITS models, the accuracy of evaluating the long-term impacts varied significantly between the two control groups. When *Y*(*t*)*c*_1_ was selected as the control group, the CITS exhibited better accuracy. When we chose *Y*(*t*)*c*_2_ as the control group, the optimized models were not as good as the classic models. The overall mean relative error was 8.68% when *Y*(*t*)*c*_1_ was chosen as the control group, whereas it was 27.62% when we chose *Y*(*t*)*c*_2_. Especially for the model CITS-segmented regression, its mean relative error with control group *Y*(*t*)*c*_1_ was 4.65%, while it became 38.53% with *Y*(*t*)*c*_2_, so the disparity between them was 33.88%. The error performance for each CITS model (CITS, CITS-A, and CITS-HA) with the two control groups is presented in **Supplementary materials Fig S7**.

#### 3.1.4 Different nonlinear orders of Hammerstein nonlinear module

In **Fig 4**, we observed that the relative error did not decrease significantly with an increase in the nonlinear order. This would increase or remain stable. Overall, the relative error was very similar, reaching its minimum in the second or third order.

**Figure 4.**
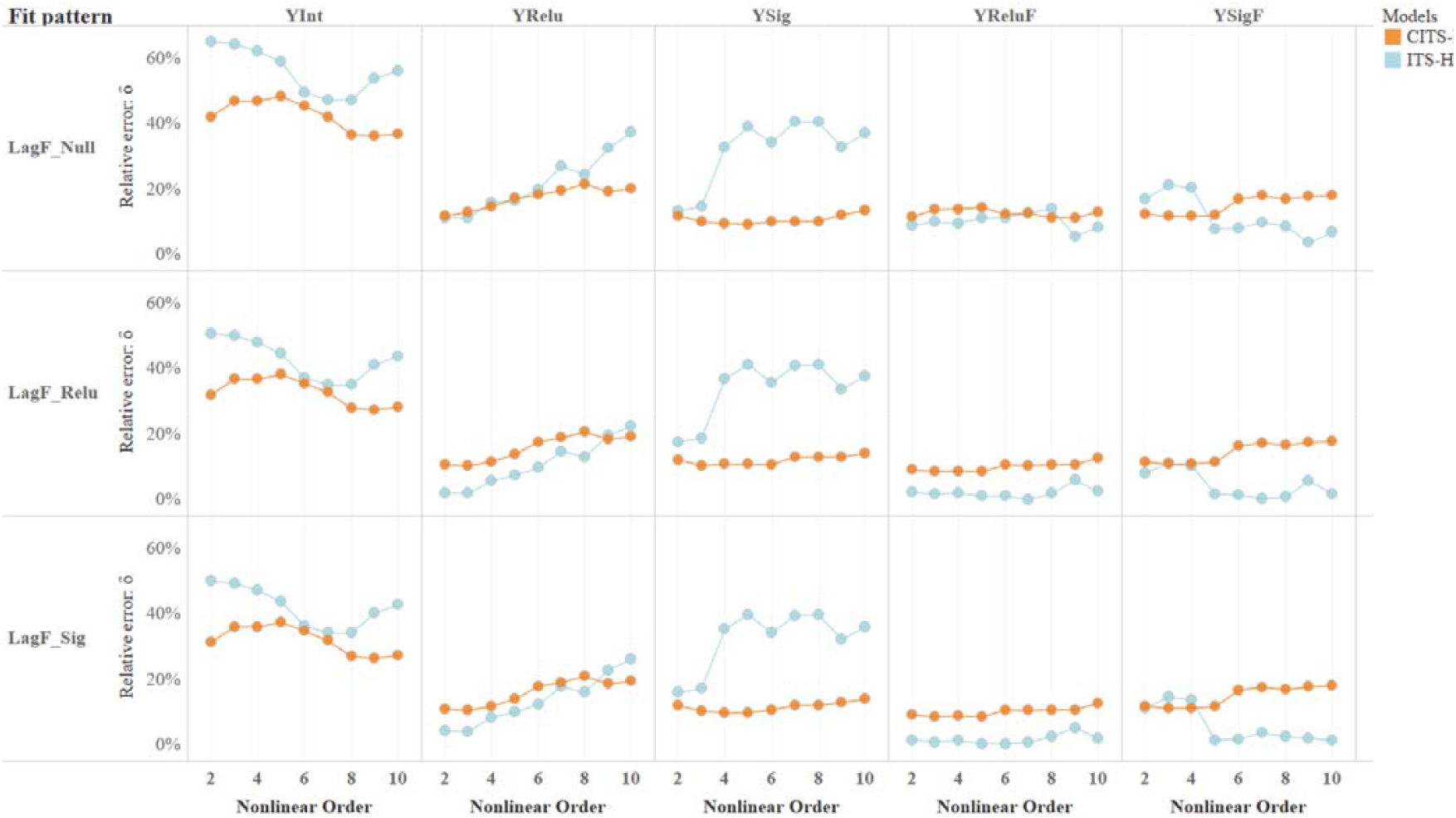
The relative error of point estimates in long-term impacts with different nonlinear orders. The horizontal axis represents the nonlinear order of the model. The vertical axis represents the relative error percentage. We considered three patterns of lag and five types of outcome series. The six colors represent the six different models. Each dot indicates the mean relative error (δ) in a fixed order.

### 3.2 Precision of evaluation

#### 3.2.1 Optimized models VS. Classic segmented regression models

Overall, there were considerable differences in the width of the 95% CI among the various models, with the widest in the ITS-HA model (0.1633) and the narrowest in the ITS model (0.0758) (**Table 3, Fig S8**). We noted that the 95% CI of 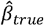 was wider in the optimized models than in the classic segmented regression models. The average width of the 95% CI in the optimized models was up to 0.1261, compared with 0.0875 in the classic model. The mean width of the four optimized models was 0.1261, compared to 0.0875 for the classic model (ITS 0.0758 to ITS-A 0.1393 and ITS-HA 0.1633 for the ITS model class; CITS 0.0992 to CITS-A 0.1245 and CITS-HA 0.1284 for the CITS model class), and the width of the 95% CI in the optimized models was expanded by approximately 58.71%. The results were similar in terms of other confidence levels (CL) (*CL* = 90% − 99.9%, *step* = 0.1%) (**Fig. S9**).

**Table 3.** Width of 95% CI of 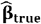 for different models.

| Model | Control group | LagF_Null | LagF_Relu | LagF_Sig | Mean width of 95% CI | Coverage rate (%) |
| --- | --- | --- | --- | --- | --- | --- |
| ITS | - | 0.0773 | 0.0749 | 0.0752 | 0.0758 | 73.33 |
| ITS-A | - | 0.1433 | 0.1370 | 0.1375 | 0.1393 | 100 |
| ITS-HA | - | 0.1691 | 0.1600 | 0.1607 | 0.1633 | 100 |
| CITS | $Y(t)c_1$ | 0.0974 | 0.0940 | 0.0945 | 0.0953 | 100 |
| | $Y(t)c_2$ | 0.1054 | 0.1017 | 0.1022 | 0.1031 | 66.67 |
|  | Mean | 0.1014 | 0.0978 | 0.0983 | 0.0992 | 83.33 |
| CITS-A | $Y(t)c_1$ | 0.1154 | 0.1107 | 0.1112 | 0.1124 | 100 |
| | $Y(t)c_2$ | 0.1404 | 0.1343 | 0.1350 | 0.1365 | 100 |
|  | Mean | 0.1279 | 0.1225 | 0.1231 | 0.1245 | 100 |
| CITS-HA | $Y(t)c_1$ | 0.1206 | 0.1154 | 0.1160 | 0.1173 | 100 |
| | $Y(t)c_2$ | 0.1434 | 0.1372 | 0.1378 | 0.1395 | 100 |
|  | Mean | 0.1320 | 0.1263 | 0.1269 | 0.1284 | 100 |

Note: “-” means data because there was no control group for the ITS model class (ITS, ITS-A, ITS-HA). LagF\_Null (%), LagF\_Relu (%), and LagF\_Sig (%) showed a width of the 95% CI of $\hat{\beta}_{true}$ when considering three lagged patterns: no lag, linear lag, and nonlinear lag of intervention effects, respectively. The coverage rate (%) was the true value coverage percentage of the corresponding 95% CI.

#### 3.2.2 CITS model class VS. ITS model class

For the ITS and CITS model classes, the mean widths of 95% CI were 0.1174 and 0.1261, respectively, which were quite close. Moreover, the values did not change significantly when selecting distinct outcome series or different nonlinear orders (**Fig S8)**.

#### 3.2.3 CITS model class with Y(t)c_l_ VS. CITS model class with *Y*(*t*)*c*_2_

Similar to the results of the accuracy assessment, for the CITS model class, the 95% CI width varied with the selection of the control group. The mean width was 0.1149 when choosing *Y*(*t*)*c*_l_, which was slightly lower than that of *Y*(*t*)*c*_2_ (0.1359).

#### 3.2.4 Coverage rate of 95% CI

As shown in **Fig S10-S15**, the 95% CI of 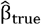 in the optimized models covered the true values *β*_*true*_ in all evaluation scenarios, whereas the 95% CIs of the classic ITS and CITS models covered the true values by 73.33% (11/15) and 83.33% (25/30), respectively.

## 4. Discussion

In this study, to overcome the defects of classic segmented regression models and to analyze the nonlinear effects of covariates on the outcome series, we proposed four optimized models within the analytical framework of ITS/CITS designs: ITS-A, ITS-HA, CITS-A, and CITS-HA. At the same time, we evaluated the accuracy and precision of evaluating the long-term impacts of the intervention for four optimized models and two classic segmented regression models based on simulated data. Overall, the optimized models were more accurate in evaluating the intervention effect. Specifically, the mean relative errors in ITS-A (14.34%) and ITS-HA (21.47%) were lower than those in the classic ITS model (26.66%), and the mean relative errors in the optimized CITS-A (16.57%) and CITS-HA (17.94%) were lower than those in the classic CITS model (21.59%).

Compared with the ITS, the CITS design, in theory, can minimize bias caused by unmeasured factors due to co-interventions or other events occurring during the intervention period by adding a control series. Therefore, both a before-after comparison and an intervention-control group comparison were considered. On average, we found that the mean relative error in the CITS model class was significantly lower than that in the ITS model class. This phenomenon was not only observed in the classic segmented regression models but also in their extended models (CITS-HA relative to ITS-HA), which was demonstrated in line with previous research on the efficacy of CITS design to minimize potential bias arising from unmeasured factors from simultaneous events.[31]

However, it should be noted that for the CITS model class, the accuracy of the intervention evaluation varied remarkably with the selection of the control group. The mean relative error with *Y*(*t*)*c*_2_ (control group 2) was significantly higher than that with *Y*(*t*)*c*_l_ (control group 1) and even higher than that in the ITS model class in some scenarios. This indicates that possible unmeasured factors in the control group or the inclusion of inappropriate covariates could have a remarkably negative impact on the accuracy of intervention evaluation in the CITS model class, and an inappropriate choice of the control may reduce the validity of the design.[32] However, little research is available on the selection of an appropriate control for CITS design. Unlike randomized controlled trials, in which a control group is obtained through randomization, in CITS design, a control series is usually selected based on research experience. Bernal et al. summarized the six controls that are most commonly used: location-based controls, characteristic-based controls, behavior-based controls, and historical cohort controls. However, each method has its own strengths and limitations.[31] Our results suggest that when there is no existing control series or a suitable control series cannot be identified, we can either choose the ITS model directly or assign weights to a pool of eligible controls using a synthetic control methodology.[33–35] Ariel proposed a framework that combined synthetic control by identifying a combination of non-intervened groups to serve as a plausible control group, while capitalizing on the CITS to provide robust impact estimates.[35] However, synthetic controls also rely on the availability of multiple suitable controls with various measures based on several characteristics and may not completely eliminate significant validity risks in an interrupted time-series analysis.[33] The performance of synthetic controls in CITS studies needs to be studied further.

With respect to precision, the width of the CI of intervention effects was larger in the optimized models than in the classic models, regardless of confidence levels. The optimized models consider extra uncertainty about the covariate and intervention series (nonlinear effects and autocorrelation), which correspondingly widen the width of the CI. In this study, such extra considerations of uncertainty were supported by the fact that the 95% CI of optimized models covered true values in all scenarios, whereas classic models did not (coverage rate:73.33% for ITS and 83.33% for CITS). It should be noted that the rate of true values covered by the corresponding CI is more important for understanding the parameter distributions.[36, 37] We argue that the right method should be the one that can minimize the standard error on the premise that the parameter is still included in the CI for impact estimation.

In addition, in the models (ITS-HA and CITS-HA) with a nonlinear structure, there was no significant drop in the relative error with the nonlinear order of covariates. This indicates that, in the actual process, a higher nonlinear order does not necessarily yield better results, and the interference of noise in the covariate may also increase with the increase in nonlinear orders.[38, 39] Therefore, when we considered the nonlinear effects of covariates, higher orders did not indicate a better model performance. In our simulation analyses, no more than three orders would be appropriate.

### 4.1 Strengths

The disentanglement of the nonlinear effects of covariates was a key strength and a novel aspect of this study. To control for the nonlinear effects of covariates, the embedding Hammerstein model can transform the control problem of nonlinear systems into linear systems with simple structures. It can effectively describe nonlinear dynamic systems[40] and has better interpretability than other nonlinear models (e.g., grey-box models and neural networks).[41]

In this process, to reflect the actual intervention time-series characteristics, we constructed a sequence generator for simulation testing, which provided an approach to generating time series with particular characteristics rather than the simple and repetitive Monte Carlo simulation,[42, 43] which was useful for further data simulation studies. At the same time, we comprehensively analyzed different model performances using different outcome series, lagged patterns of intervention effect, nonlinear orders, and control groups for the CITS model class, 495 different combinations altogether, which had a comprehensive description of interrupt time series analysis under various circumstances and provided a reference for selecting nonlinear order in modeling.

### 4.2 limitations

This study had some limitations. First, for the advantages and disadvantages of different models, we only compared the relative error of their point estimates and focused on long-term impacts, as did the precision assessment. However, we did not consider the robustness of point estimation or differences in the statistical power of the corresponding hypothesis testing among different models [43, 44]. Second, to estimate long-term impacts, we used ordinary least squares (OLS) as the estimation algorithm. There are other algorithms in parameter estimation, such as generalized least squares and restricted maximum likelihood, which are not exactly consistent across algorithms.[45, 46] Thirdly, we only considered the autocorrelation of the outcome series itself, without considering the autocorrelation of the error terms, i.e., the ‘*MA*’ part of the ARIMAX model which was necessary and valuable in some situations.[47–49] Fourth, the optimized models we proposed by insetting Hammerstein require a strong correlation between covariate sequence and outcome series, in which case researchers should be prudential about the inclusion of covariates. This explains why we stimulated the outcome series through a nonlinear transformation of the covariates. However, in practical applications, it is challenging to identify a good covariate series.

## 5. Conclusion

The ITS/CITS-segmented regression models have been powerful for evaluating intervention effectiveness, and their use has been increasing. They are not always adequate, especially when there may be nonlinear effects of the covariates. Based on the simulated data, the optimized models (ITS/CITS-A, ITS/CITS-HA) performed better than the classic models (ITS/CITS) in terms of accuracy, however they were not as precise as the classic models in terms of interval widths regardless of confidence level. The optimized models are useful tools as they can assess the long-term impact of intervention with additional considerations for covariates’ nonlinear effects and allow for modeling of time-series autocorrelation and lag of intervention effects.

## Data Availability

All data produced in the present study are available upon reasonable request to the authors

## Ethics approval and consent to participate

Not applicable.

## Consent for publication

Not applicable.

## Availability of data and materials

The datasets used and/or analyzed during the current study are available from the corresponding author on reasonable request.

## Authors’ contributions

XL Zhang designed the study and performed data analyses. XL Zhang and R Yin wrote the first draft and developed subsequent manuscript. To be more specific, XL Zhang is in the charge of conceptualization, data curation, methodology, formal analysis, software, visualization and writing-original draft. R Yin is responsible for supervision, validation and writing both for original draft and review & editing. Meanwhile, XL Zhang and R Yin have directly accessed and verified the underlying data reported in the manuscript together. Y Pan, WF Zhong and D Kong participated in the writing and revision of the paper, and gave some constructive comments. As the corresponding author, W Chen participate all the above processes, guided the writing and critically revised the manuscript. All authors read and approved the final manuscript.

## Acknowledgements

Not applicable.

## Supplementary materials

**Figure S1.**
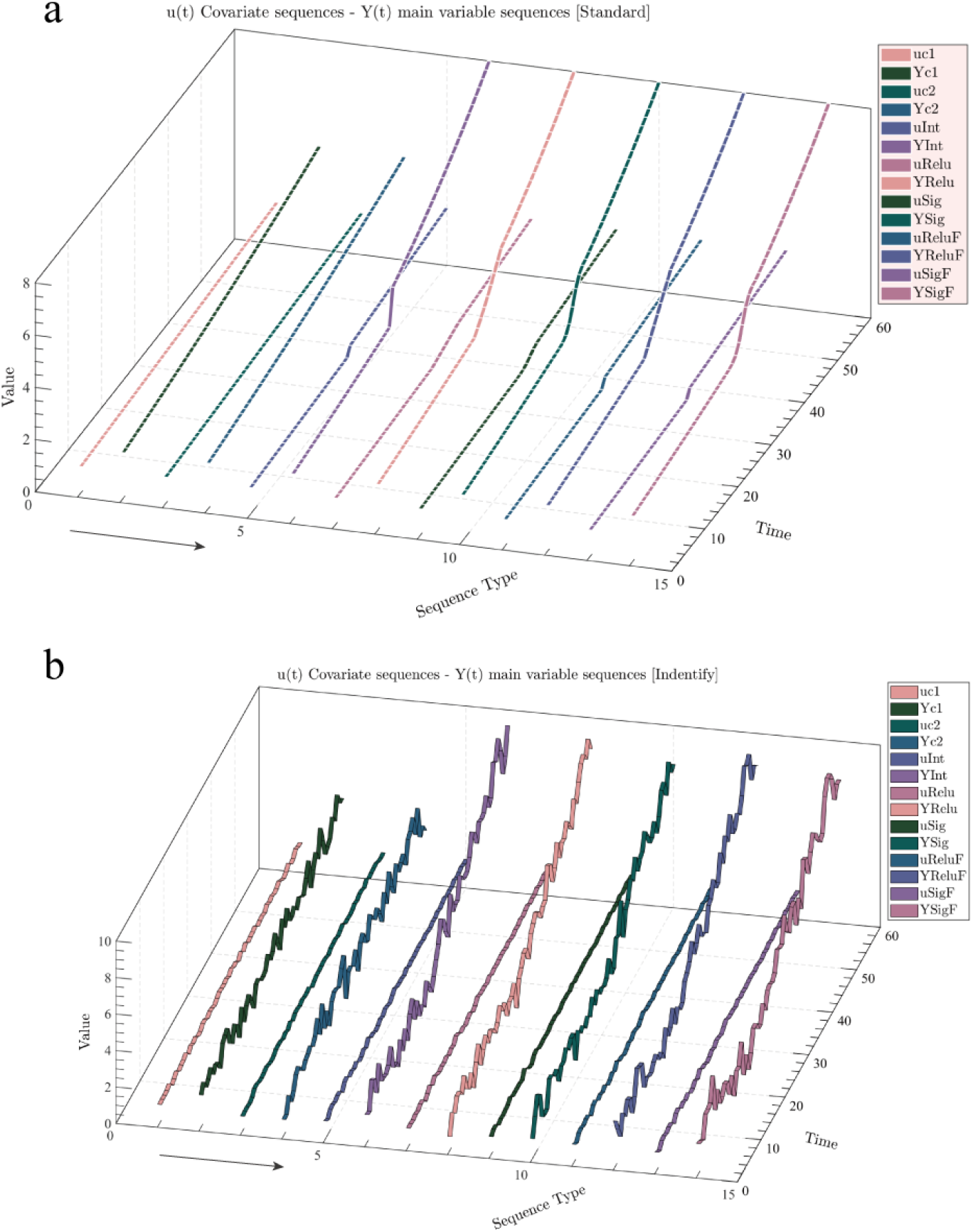
Outcome series generation consequences display chart. The difference between panel a) and panel b) lies in the presence or absence of noise entry. The series in panel a) have no noise entry and present the main structure of the sequence generation. The series in panel b) both have the entry of noise in the generation of the covariate sequence and the outcome series generation process. The order of presentation of the sequences of the two plots is the same, both are covariate sequences first, followed by their corresponding outcome series, and then alternately presented in sequence. The order is:

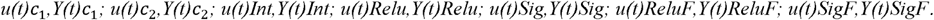

**Figure S2.**
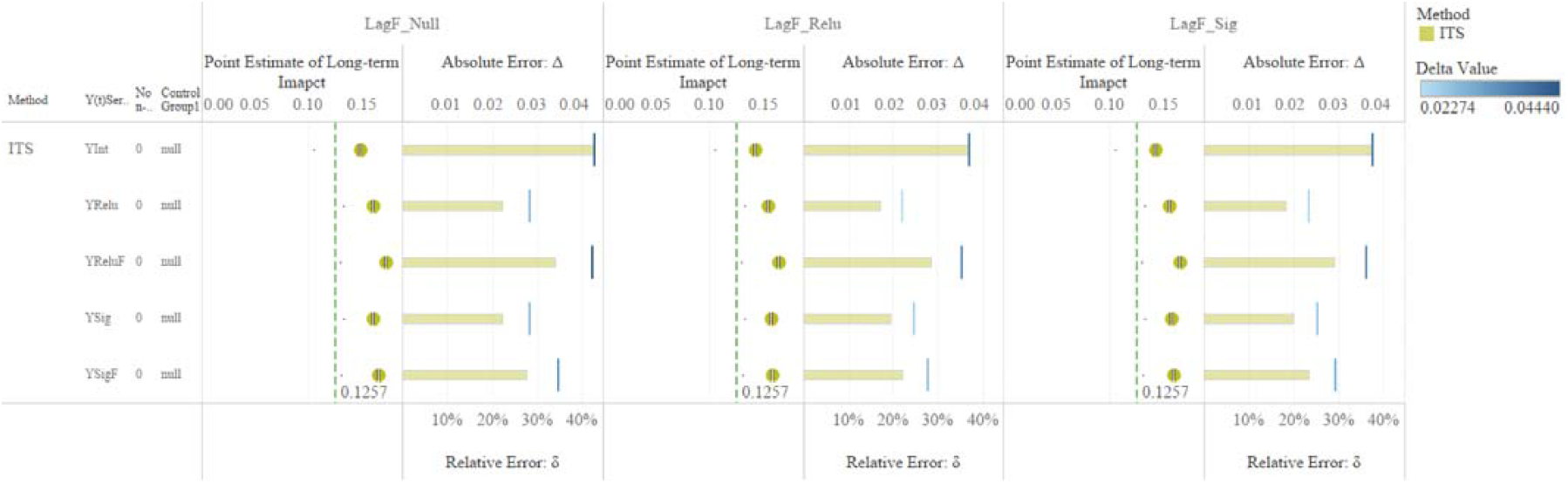
Point estimates of long-term impact based on the ITS model and their corresponding error rates. The vertical columns represent three lag patterns: no lag (LagF_Null), linear lag (LagF_Relu), and nonlinear lag (LagF_Sig). Each column is divided into two sub-vertical columns. In the left sub-vertical column, the dots represent the point estimate of the long-term impacts of an evaluation based on ITS. The green, thin, and dashed lines represent the mean of the true values of the long-term impacts. In the right sub-vertical column, the height of each bar represents the magnitude of the relative error δ, and the broken vertical lines represent the absolute error Δ.

**Figure S3.**
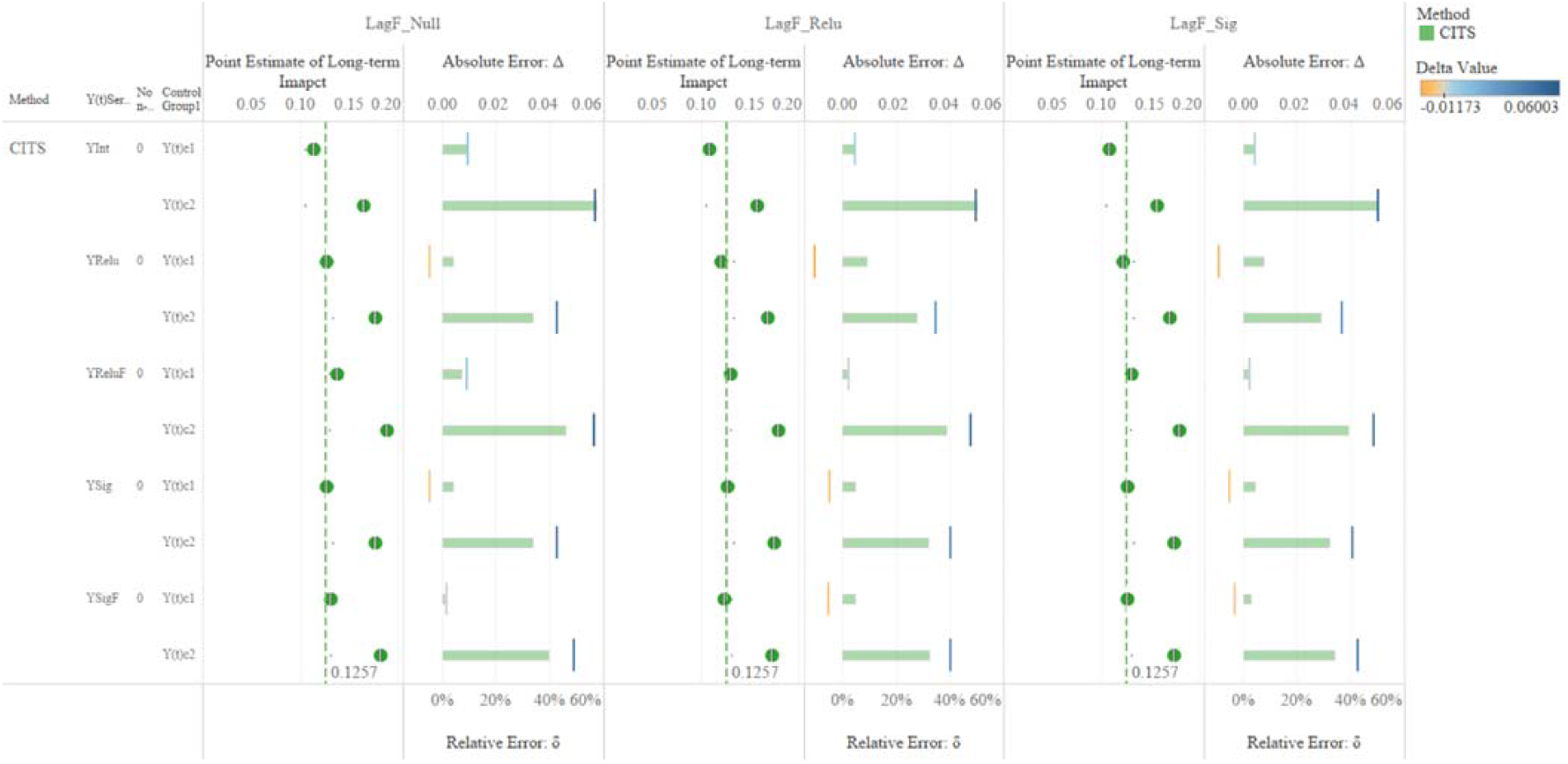
Point estimates of long-term impact based on the CITS model and their corresponding error rates. The vertical columns represent three lag patterns: no lag (LagF_Null), linear lag (LagF_Relu), and nonlinear lag (LagF_Sig). Each column is divided into two sub-vertical columns. In the left sub-vertical column, the dots represent the point estimate of the long-term impacts of an evaluation based on CITS. The green, thin, and dashed lines represent the mean of the true values of the long-term impacts. In the right sub-vertical column, the height of each bar represents the magnitude of the relative error δ, and the broken vertical lines represent the absolute error Δ.

**Figure S4.**
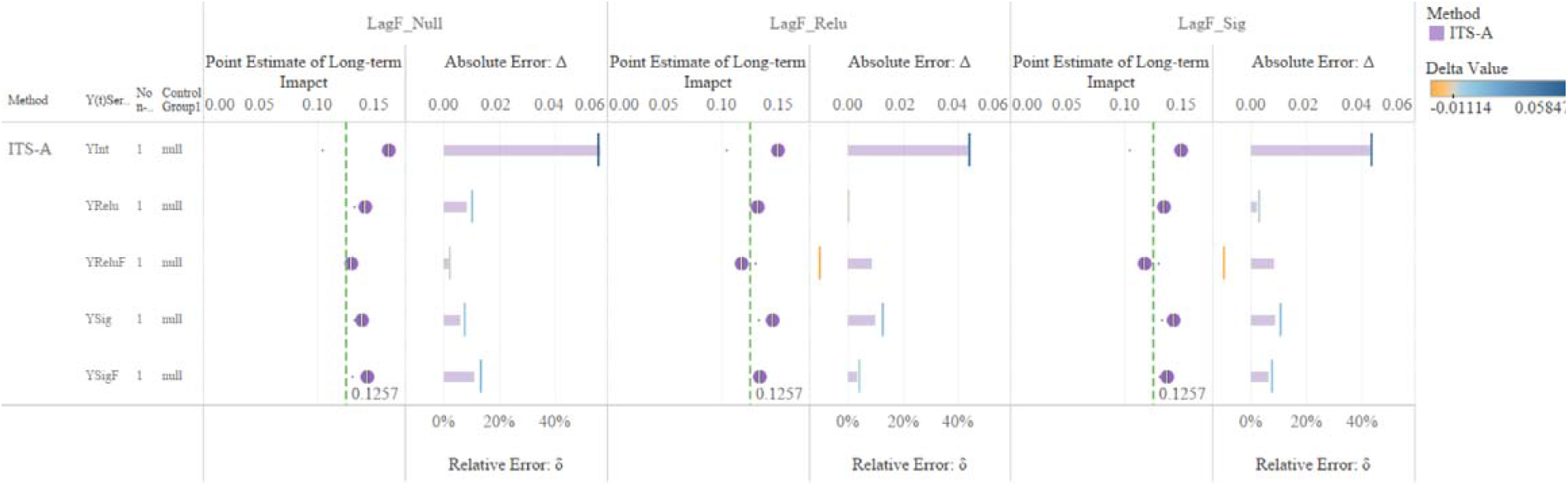
Point estimates of long-term impact based on the ITS-A model and their corresponding error rates. The vertical columns represent three lag patterns: no lag (LagF_Null), linear lag (LagF_Relu), and nonlinear lag (LagF_Sig). Each column is divided into two sub-vertical columns. In the left sub-vertical column, the dots represent the point estimate of the long-term impacts of an evaluation based on ITS-A. The green, thin, and dashed lines represent the mean of the true values of the long-term impacts. In the right sub-vertical column, the height of each bar represents the magnitude of the relative error δ, and the broken vertical lines represent the absolute error Δ.

**Figure S5.**
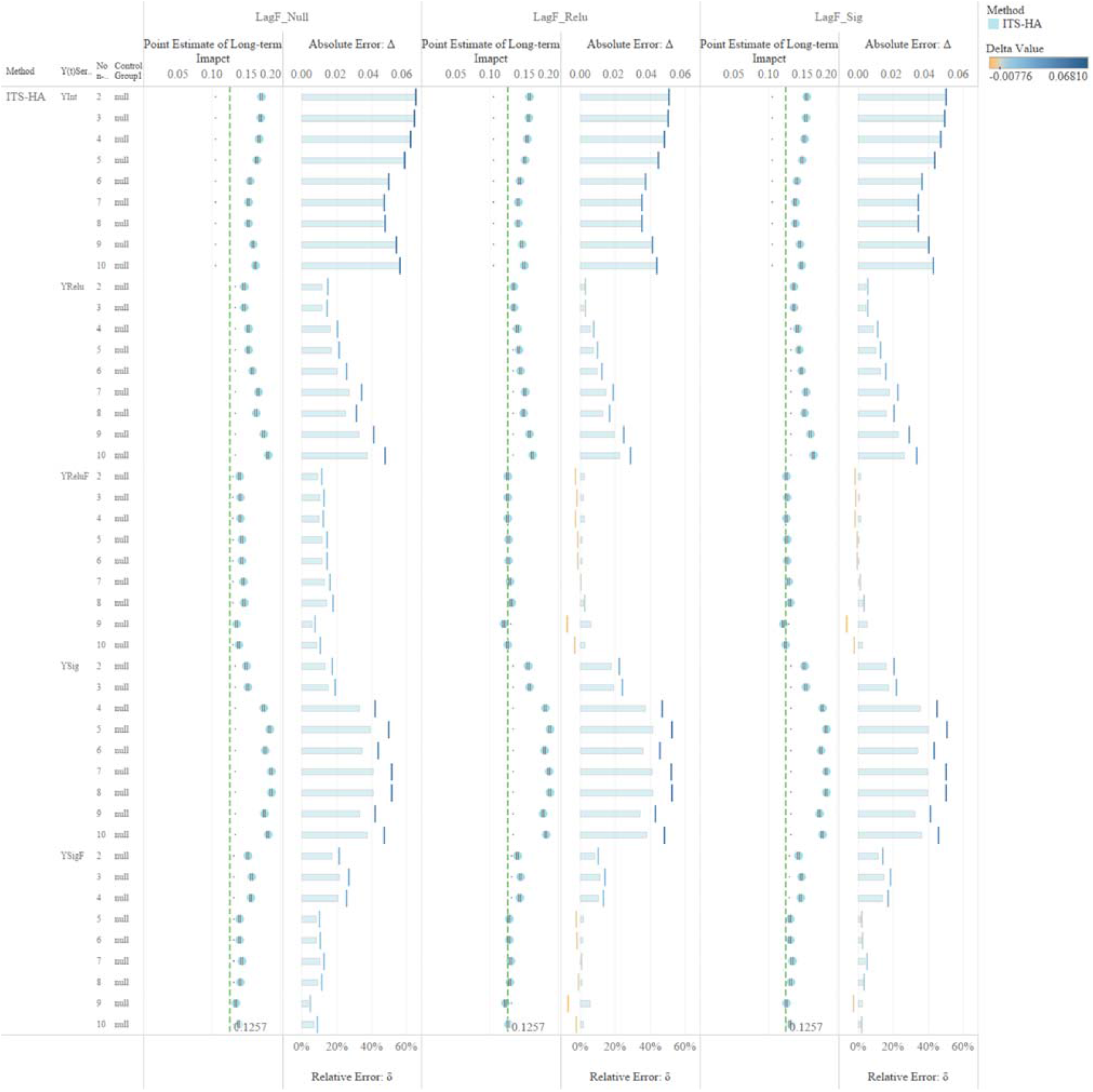
Point estimates of long-term impact based on the ITS-HA model and their corresponding error rates. The vertical columns represent three lag patterns: no lag (LagF_Null), linear lag (LagF_Relu), and nonlinear lag (LagF_Sig). Each column is divided into two sub-vertical columns. In the left sub-vertical column, the dots represent the point estimate of the long-term impacts of an evaluation based on ITS-HA. The green, thin, and dashed lines represent the mean of the true values of the long-term impacts. In the right sub-vertical column, the height of each bar represents the magnitude of the relative error δ, and the broken vertical lines represent the absolute error Δ.

**Figure S6.**
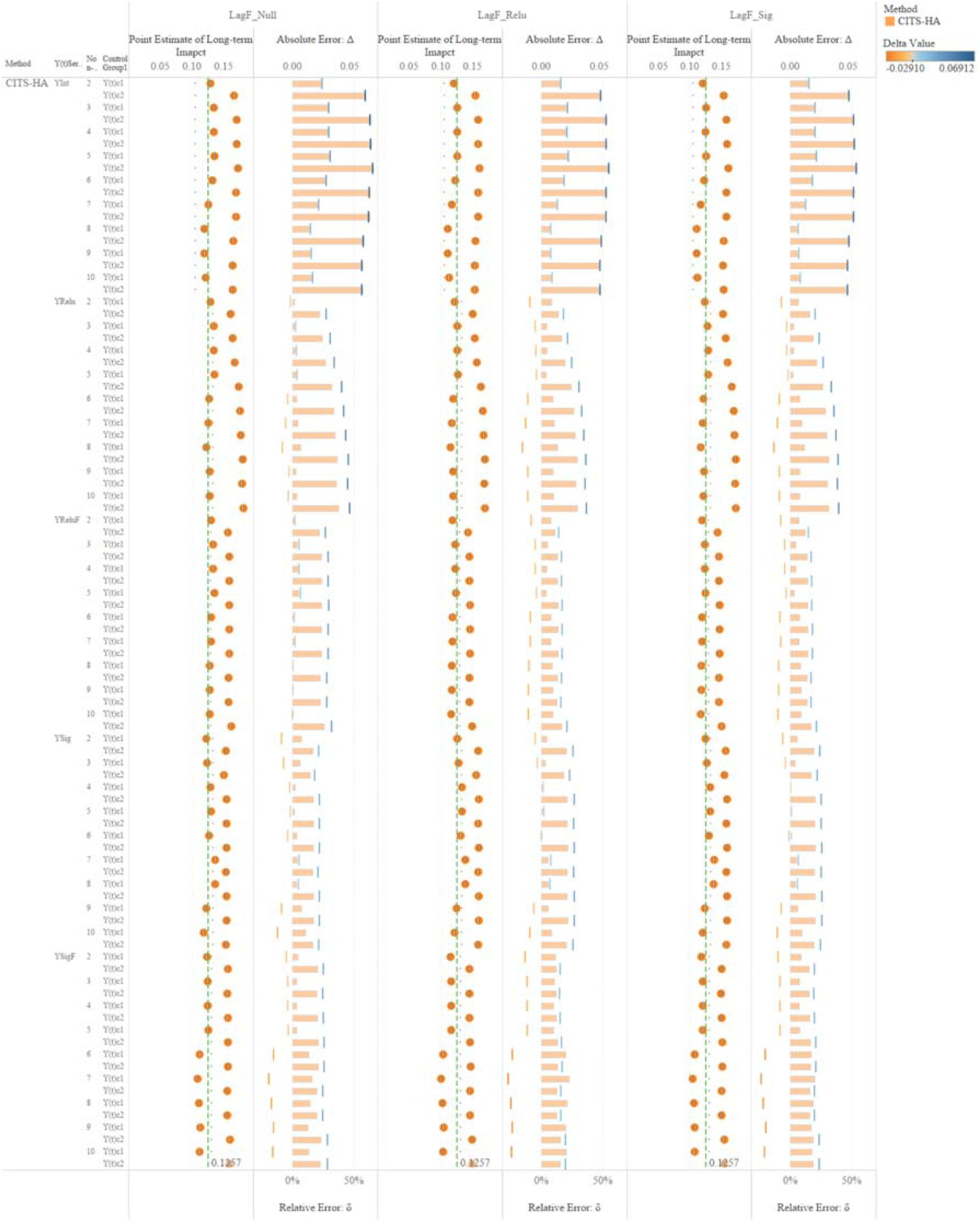
Point estimates of long-term impact based on the CITS-HA model and their corresponding error rates. The vertical columns represent three lag patterns: no lag (LagF_Null), linear lag (LagF_Relu), and nonlinear lag (LagF_Sig). Each column is divided into two sub-vertical columns. In the left sub-vertical column, the dots represent the point estimate of the long-term impacts of an evaluation based on CITS-HA. The green, thin, and dashed lines represent the mean of the true values of the long-term impacts. In the right sub-vertical column, the height of each bar represents the magnitude of the relative error δ, and the broken vertical lines represent the absolute error Δ.

**Figure S7.**
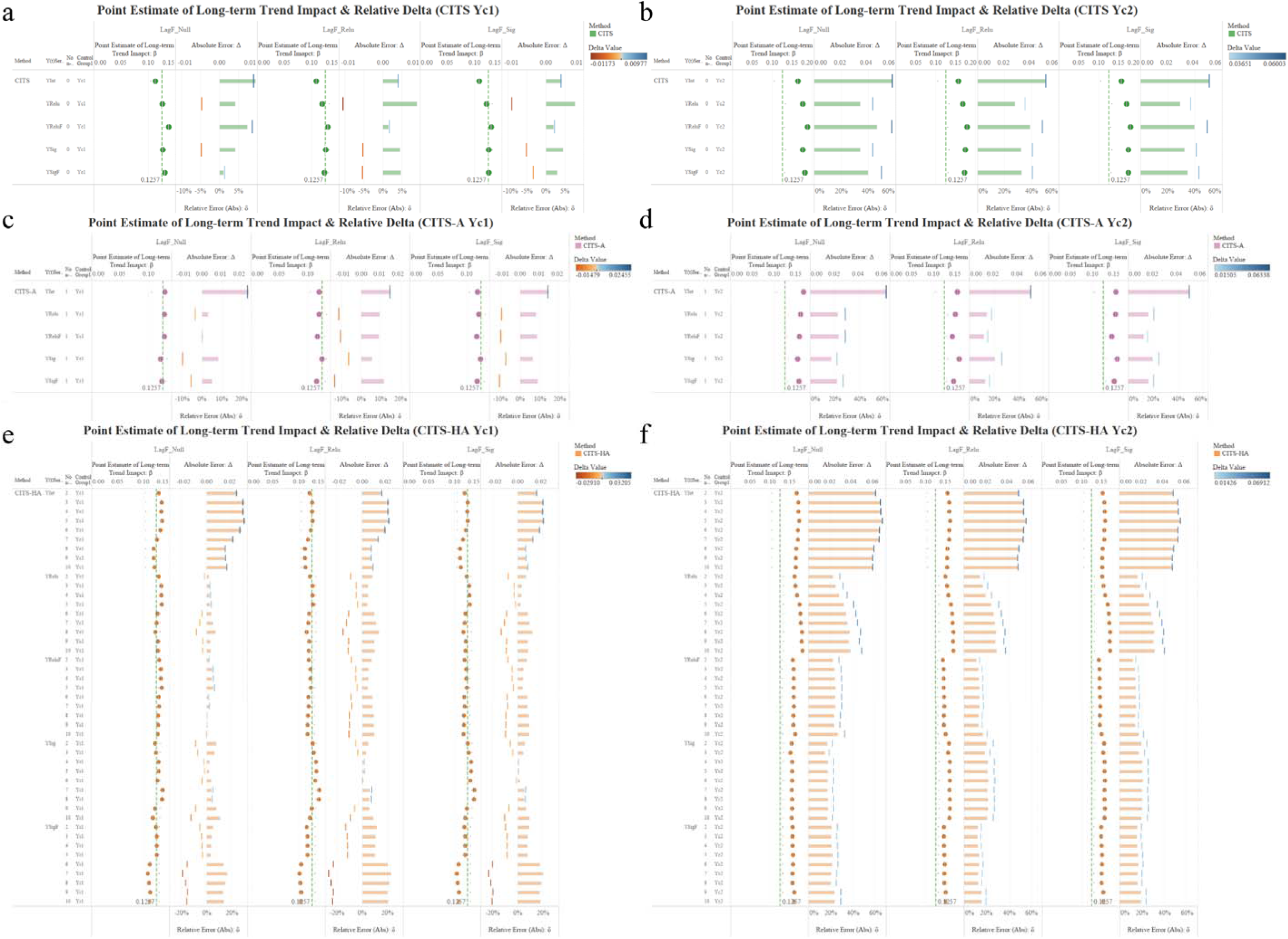
Point estimation of long-term impact and their corresponding error rates with two control groups.

**Figure S8.**
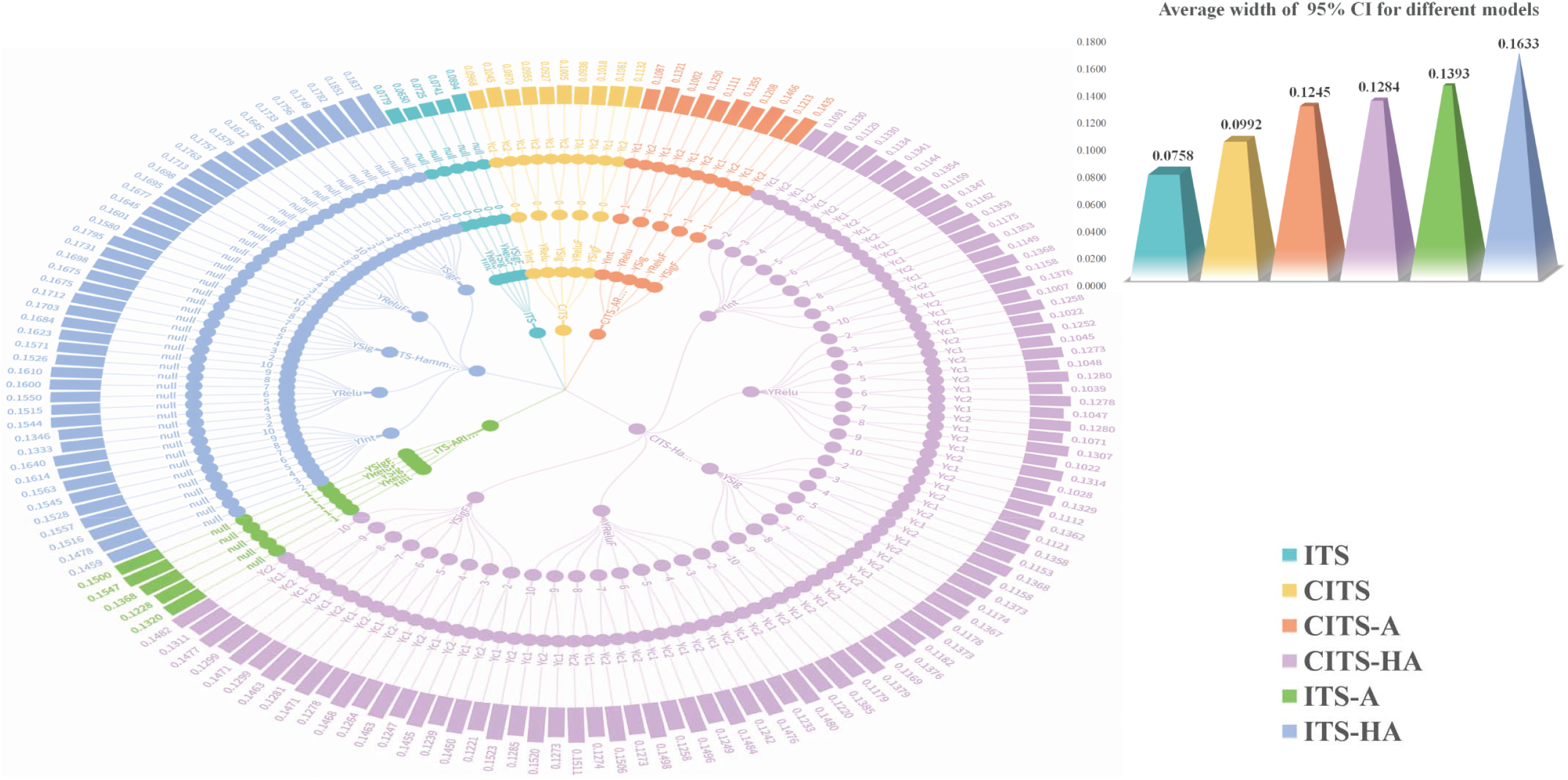
95% CI widths of different models. Different colors represent different models. The main radical figure is divided into a total of five layers from inside to outside, with the first layer representing different models, the second layer representing different sequences, the third layer representing different nonlinear orders, and the fourth layer representing different control groups. The outermost circled level, the fifth, with different bar heights represents the size of 95% CI width of long-term impact. The heights of the prisms of the smaller graph which is in the upper right corner of the main figure, then, represent the average value of the long-term trend 95% CI width for different models.

**Figure S9.**
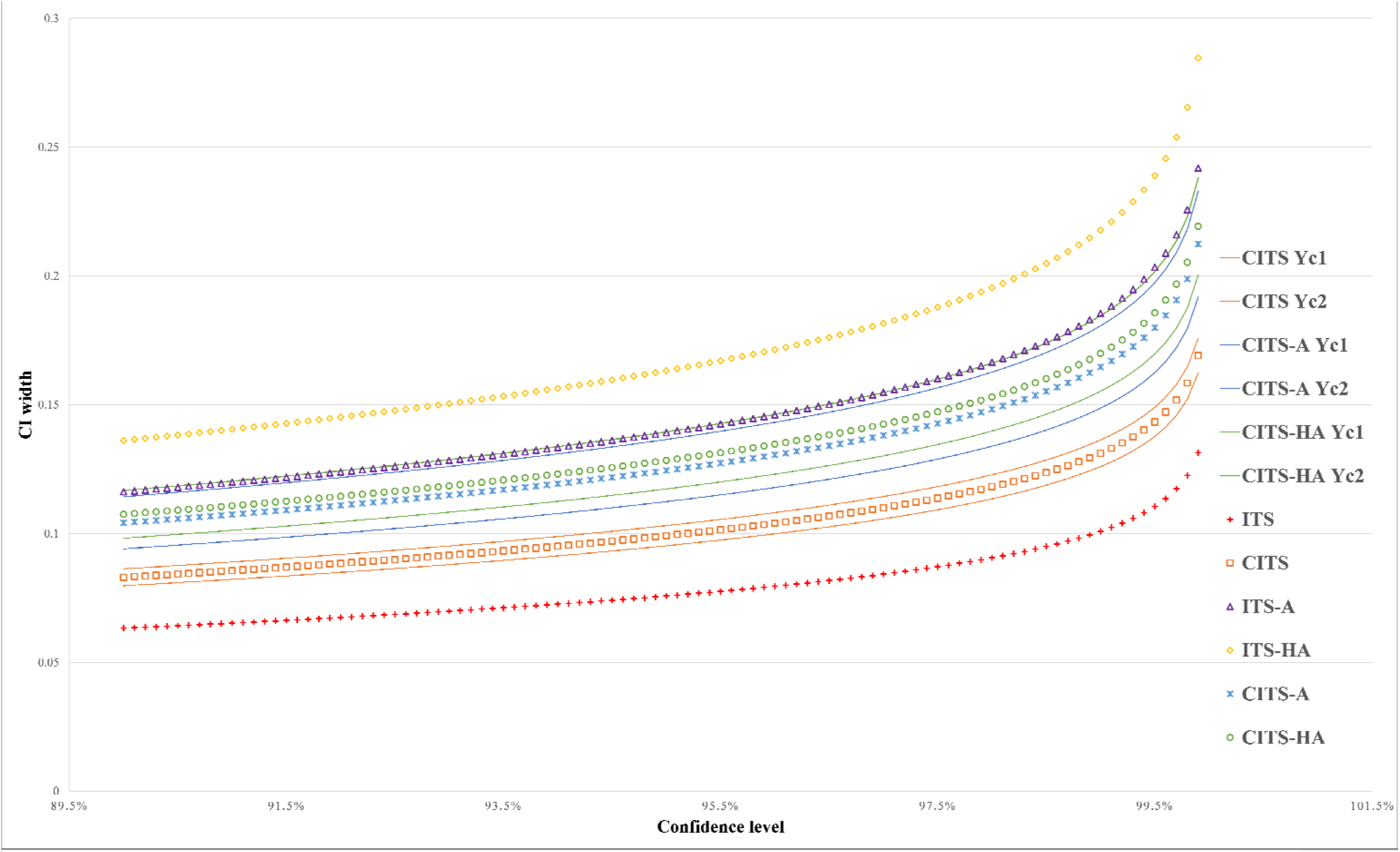
CI widths under different confidence levels.

**Figure S10.**
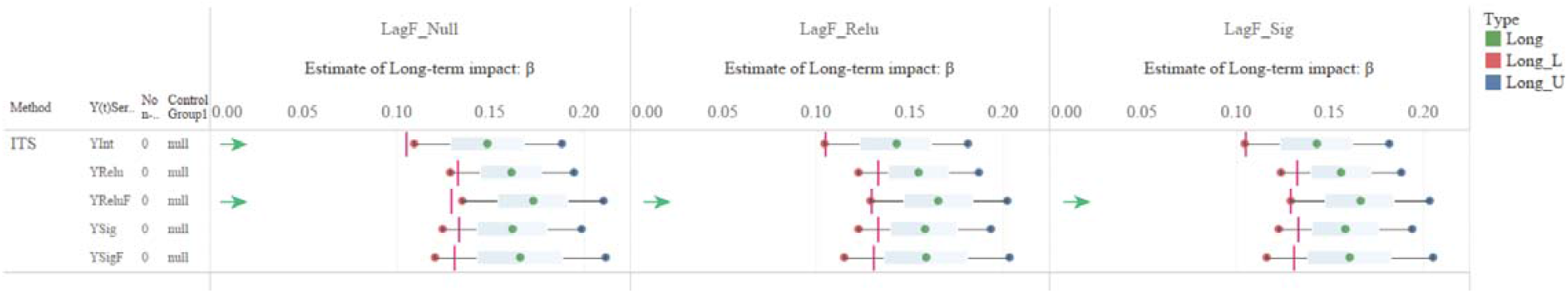
95% confidence interval of long-term impact with ITS. The small red vertical bars represent the true values of the long-term impact of the different outcome series. The left and right ends of the box-and-whisker plot represent the lower and upper boundary values of the long-term impact of 95% confidence interval, respectively. The presence of the green arrow on the left means that in this scenario, the true value is not included in the range of the 95% estimation interval.

**Figure S11.**
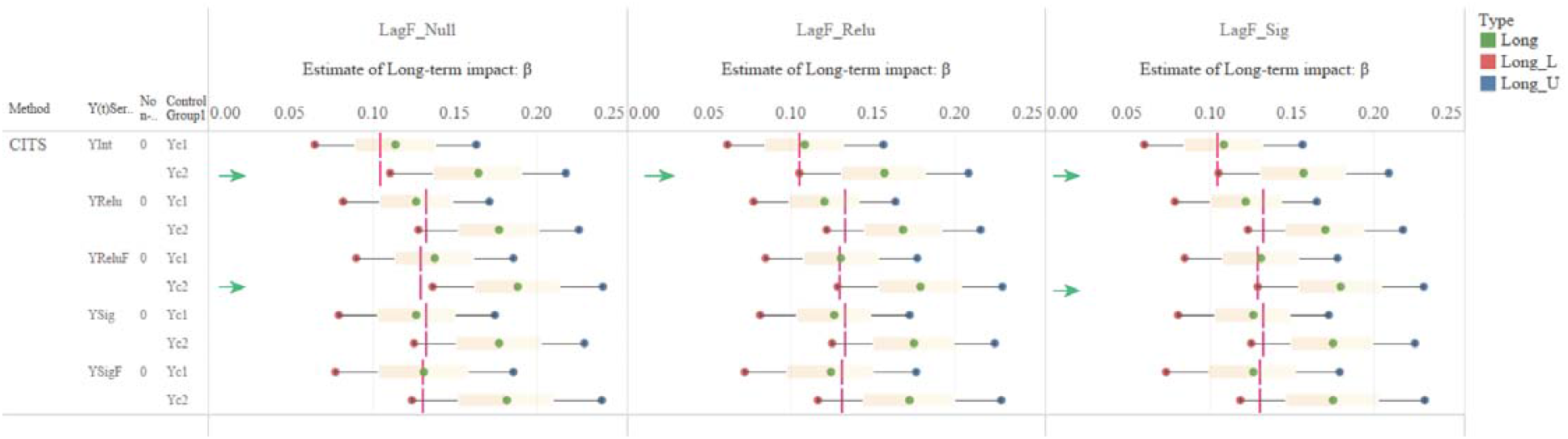
95% confidence interval of long-term impact with CITS. The small red vertical bars represent the true values of the long-term impact of the different outcome series. The left and right ends of the box-and-whisker plot represent the lower and upper boundary values of the long-term impact of 95% confidence interval, respectively. The presence of the green arrow on the left means that in this scenario, the true value is not included in the range of the 95% estimation interval.

**Figure S12.**
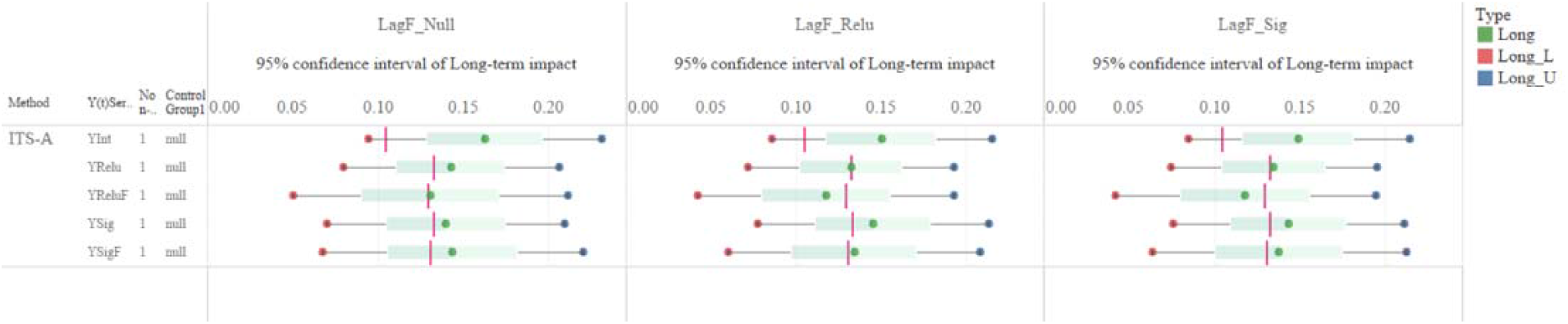
95% confidence interval of long-term impact with ITS-A. The small red vertical bars represent the true values of the long-term impact of the different outcome series. The left and right ends of the box-and-whisker plot represent the lower and upper boundary values of the long-term impact of 95% confidence interval, respectively.

**Figure S13.**
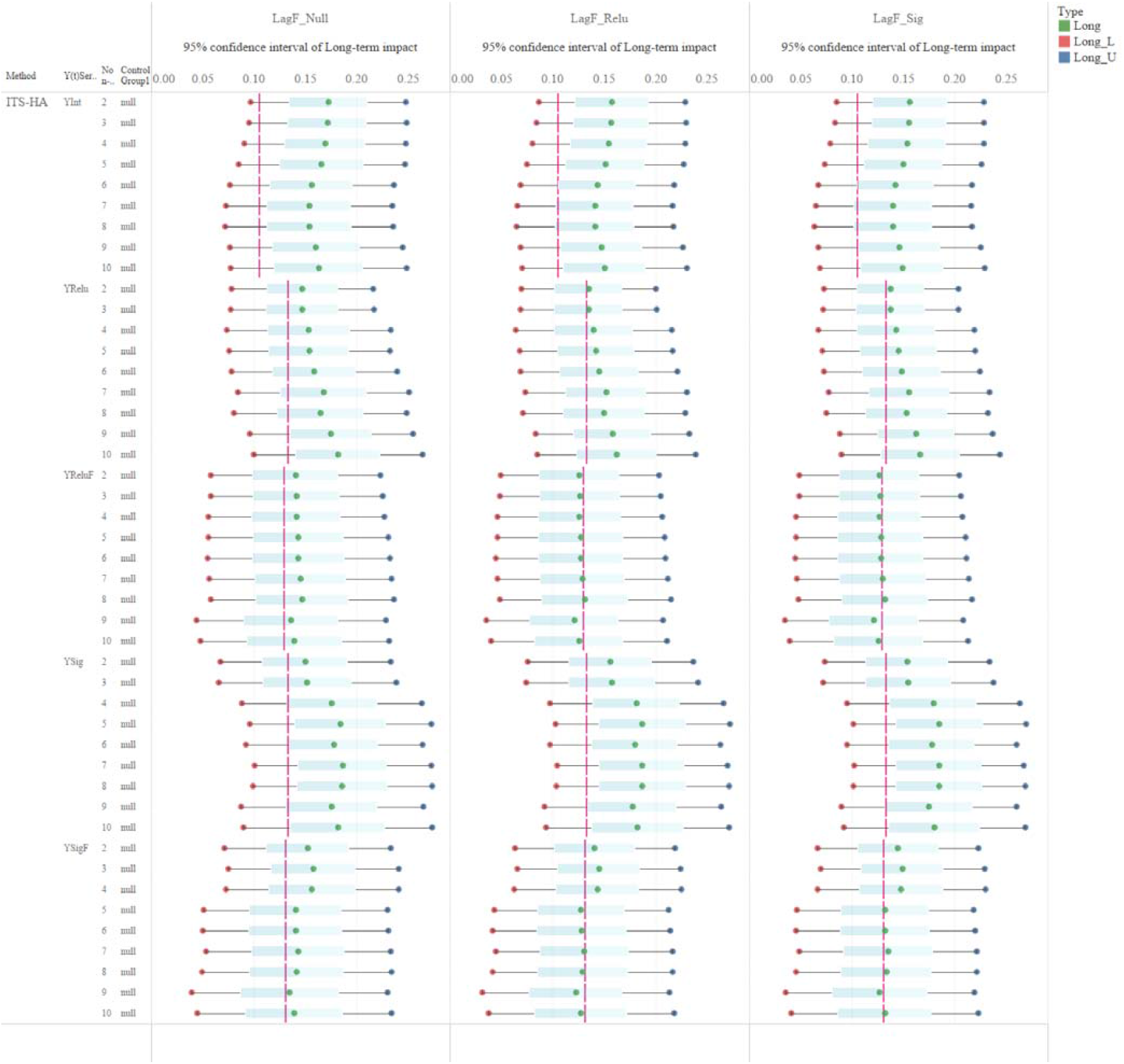
95% confidence interval of long-term impact with ITS-HA. The small red vertical bars represent the true values of the long-term impact of the different outcome series. The left and right ends of the box-and-whisker plot represent the lower and upper boundary values of the long-term impact of 95% confidence interval, respectively.

**Figure S14.**
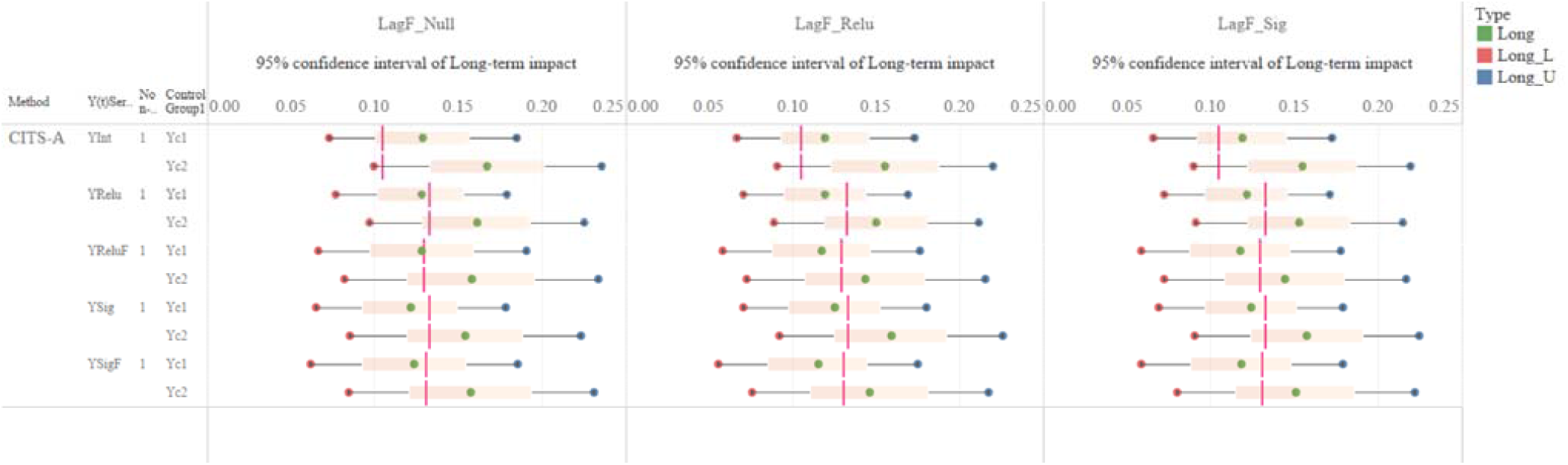
95% confidence interval of long-term impact with CITS-A. The small red vertical bars represent the true values of the long-term impact of the different outcome series. The left and right ends of the box-and-whisker plot represent the lower and upper boundary values of the long-term impact of 95% confidence interval, respectively.

**Figure S15.**
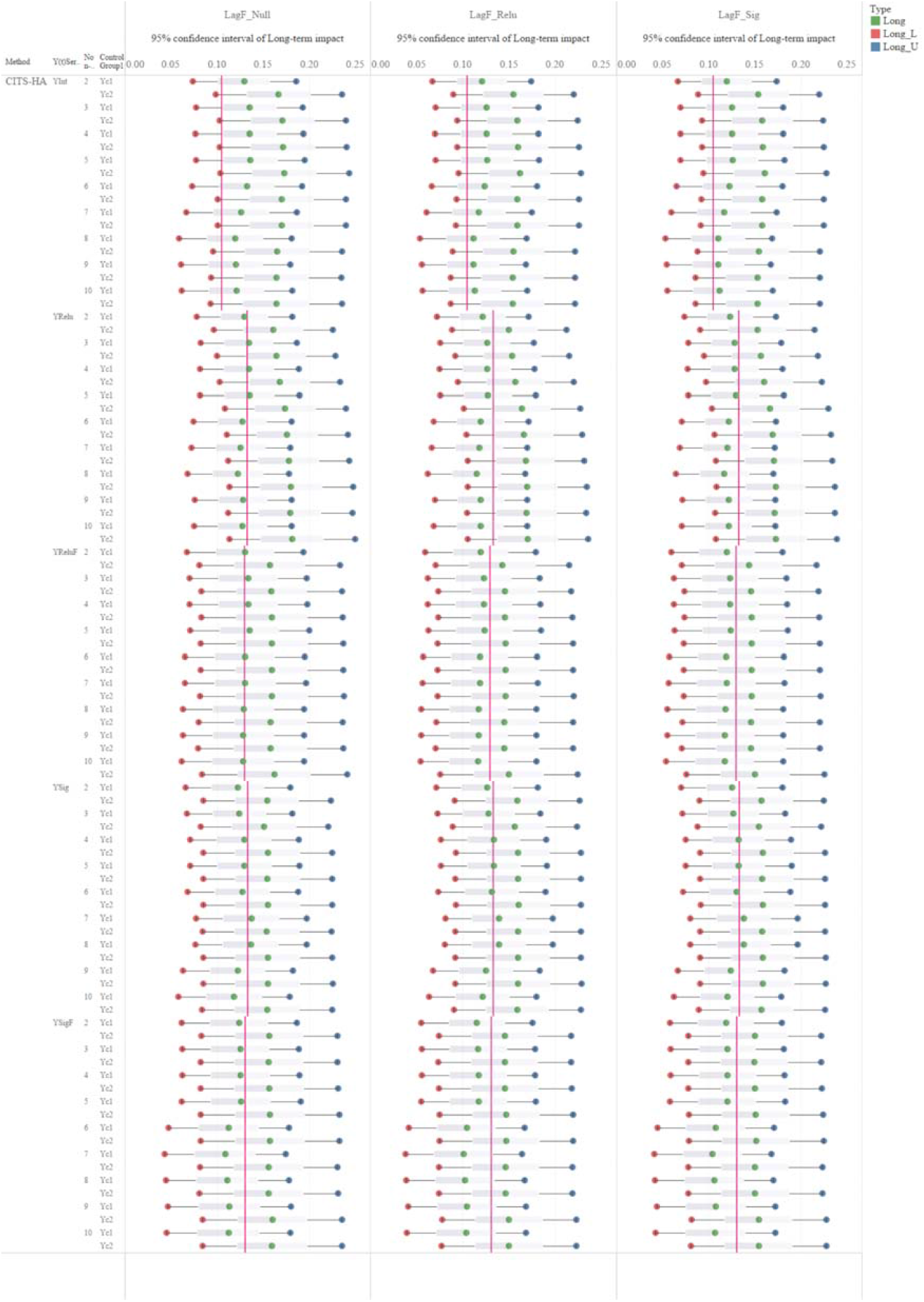
95% confidence interval of long-term impact with CITS-HA. The small red vertical bars represent the true values of the long-term impact of the different outcome series. The left and right ends of the box-and-whisker plot represent the lower and upper boundary values of the long-term impact of 95% confidence interval, respectively.

## References

1. Box GEP, Tiao GC. Intervention analysis with applications to economic and environmental problems. J Am Stat Assoc. 1975;70.

2. Harris AD, McGregor JC, Perencevich EN, Furuno JP, Zhu J, Peterson DE, et al. The Use and Interpretation of Quasi-Experimental Studies in Medical Informatics. Journal of the American Medical Informatics Association. 2006;13:16–23.

3. Lopez Bernal J, Soumerai S, Gasparrini A. A methodological framework for model selection in interrupted time series studies. J Clin Epidemiol. 2018;103:82–91.

4. Kontopantelis E, Doran T, Springate DA, Buchan I, Reeves D. Regression based quasi-experimental approach when randomisation is not an option: interrupted time series analysis. BMJ. 2015;350.

5. Wagner AK, Soumerai SB, Zhang F, Ross-Degnan D. Segmented regression analysis of interrupted time series studies in medication use research. J Clin Pharm Ther. 2002;27:299–309.

6. Schmidt AF, Finan C. Linear regression and the normality assumption. J Clin Epidemiol. 2018;98:146–51.

7. Penfold RB, Zhang F. Use of Interrupted Time Series Analysis in Evaluating Health Care Quality Improvements. Acad Pediatr. 2013;13:S38–44.

8. Hanbury A, Farley K, Thompson C, Wilson PM, Chambers D, Holmes H. Immediate versus sustained effects: interrupted time series analysis of a tailored intervention. Implementation Science. 2013;8:130.

9. Beard E, Jackson SE, West R, Kuipers MAG, Brown J. Population-level predictors of changes in success rates of smoking quit attempts in England: a time series analysis. Addiction. 2020;115.

10. Roshani D, Ghaderi E. Comparing Smoothing Techniques for Fitting the Nonlinear Effect of Covariate in Cox Models. Acta Informatica Medica. 2016;24:38.

11. Cui X, Guo W, Lin L, Zhu L. Covariate-adjusted nonlinear regression. The Annals of Statistics. 2009;37:1839–70.

12. Crainiceanu C, Ruppert D, Coresh J. COX MODELS WITH NONLINEAR EFFECT OF COVARIATES MEASURED WITH ERROR: A CASE STUDY OF CHRONIC KIDNEY DISEASE INCIDENCE. JOHNS HOPKINS UNIVERSITY, DEPT OF BIOSTATISTICS WORKING PAPERS. 2006.

13. Tzogiou C, Boes S, Brunner B. What explains the inequalities in health care utilization between immigrants and non-migrants in Switzerland? BMC Public Health. 2021;21:530.

14. Spiteri J, von Brockdorff P. Economic development and health outcomes: Evidence from cardiovascular disease mortality in Europe. Soc Sci Med. 2019;224:37–44.

15. Tang S, Shi L, Chen W, Zhao P, Zheng H, Yang B, et al. Spatiotemporal distribution and sociodemographic and socioeconomic factors associated with primary and secondary syphilis in Guangdong, China, 2005–2017. PLoS Negl Trop Dis. 2021;15:e0009621.

16. Tong H, Zhang S, Shen W, Chen H, Salazar C, Schneider A, et al. Lung Function and Short-Term Ambient Air Pollution Exposure: Differential Impacts of Omega-3 and Omega-6 Fatty Acids. Ann Am Thorac Soc. 2022;19:583–93.

17. Schwartz J. The distributed lag between air pollution and daily deaths. Epidemiology. 2000;11:320–6.

18. Ferreira Braga AL, Zanobetti A, Schwartz J. The lag structure between particulate air pollution and respiratory and cardiovascular deaths in 10 US cities. J Occup Environ Med. 2001;43:927–33.

19. Pun VC, Tian L, Yu ITS, Kioumourtzoglou MA, Qiu H. Differential distributed lag patterns of source-specific particulate matter on respiratory emergency hospitalizations. Environ Sci Technol. 2015;49:3830–8.

20. Funderburg RG, Nixon H, Boarnet MG, Ferguson G. New highways and land use change: Results from a quasi-experimental research design. Transp Res Part A Policy Pract. 2010;44:76–98.

21. Helfenstein U. The Use of Transfer Function Models, Intervention Analysis and Related Time Series Methods in Epidemiology. Int J Epidemiol. 1991;20:808–15.

22. Huitema BE, McKean JW, McKnight S. Autocorrelation Effects on Least-Squares Intervention Analysis of Short Time Series. Educational and Psychological Measurement. 2016;59:767–86.

23. Reimers S, Harvey N. Sensitivity to autocorrelation in judgmental time series forecasting. Int J Forecast. 2011;27:1196–214.

24. Yue S, Pilon P, Phinney B, Cavadias G. The influence of autocorrelation on the ability to detect trend in hydrological series. Hydrol Process. 2002;16:1807–29.

25. Helfenstein U. Box-Jenkins modelling in medical research. Stat Methods Med Res. 1996;5:3–22.

26. Jandoc R, Burden AM, Mamdani M, Lévesque LE, Cadarette SM. Interrupted time series analysis in drug utilization research is increasing: Systematic review and recommendations. In: Journal of Clinical Epidemiology. 2015.

27. Rasamoelina AD, Adjailia F, Sincak P. A Review of Activation Function for Artificial Neural Network. SAMI 2020 - IEEE 18th World Symposium on Applied Machine Intelligence and Informatics, Proceedings. 2020;:281–6.

28. Pearson RK, Pottmann M. Gray-box identification of block-oriented nonlinear models. J Process Control. 2000;10.

29. Tissaoui K. Forecasting implied volatility risk indexes: International evidence using Hammerstein-ARX approach. International Review of Financial Analysis. 2019;64:232–49.

30. Aljamaan I, Westwick D, Foley M. Prediction error identification of Hammerstein models in the presence of ARIMA disturbances. IEEE Multi-conference on Systems and Control. 2014;:403–8.

31. Bernal JL, Cummins S, Gasparrini A. The use of controls in interrupted time series studies of public health interventions. Int J Epidemiol. 2018;47.

32. McDowall D, McCleary R, Bartos BJ. Interrupted time series analysis. Oxford University Press; 2019.

33. Degli Esposti M, Spreckelsen T, Gasparrini A, Wiebe DJ, Bonander C, Yakubovich AR, et al. Can synthetic controls improve causal inference in interrupted time series evaluations of public health interventions? Int J Epidemiol. 2021;49:2010–20.

34. Kim SY, Kim H, Lee JT. Health Effects of Air-Quality Regulations in Seoul Metropolitan Area: Applying Synthetic Control Method to Controlled-Interrupted Time-Series Analysis. Atmosphere 2020, Vol 11, Page 868. 2020;11:868.

35. Linden A. Combining synthetic controls and interrupted time series analysis to improve causal inference in program evaluation. J Eval Clin Pract. 2018;24:447–53.

36. Barker RJ. Effect of Heterogeneous Survival on Bird-Banding Model Confidence Interval Coverage Rates. J Wildl Manage. 1992;56:111.

37. Christoffersen PF. Evaluating Interval Forecasts. Int Econ Rev (Philadelphia). 1998;39:841.

38. Depraetere N, Vandebroek M, Depraetere N, Vandebroek M, Vandebroek M. Order selection in finite mixtures of linear regressions. Statistical Papers 2013 55:3. 2013;55:871–911.

39. Stoica P, Selén Y. Model-order selection: A review of information criterion rules. IEEE Signal Process Mag. 2004;21:36–47.

40. Jin Q, Wang H, Su Q, Jiang B, Liu Q. A novel optimization algorithm for MIMO Hammerstein model identification under heavy-tailed noise. ISA Trans. 2018;72:77–91.

41. Hart A, Wyatt J. Evaluating black-boxes as medical decision aids: issues arising from a study of neural networks. Medical Informatics. 1990;15:229–36.

42. Morris TP, White IR, Crowther MJ. Using simulation studies to evaluate statistical methods. Stat Med. 2019;38:2074–102.

43. Zhang F, Wagner AK, Ross-Degnan D. Simulation-based power calculation for designing interrupted time series analyses of health policy interventions. J Clin Epidemiol. 2011;64:1252–61.

44. McCaw ZR, Colthurst T, Yun T, Furlotte NA, Carroll A, Alipanahi B, et al. DeepNull models non-linear covariate effects to improve phenotypic prediction and association power. Nat Commun. 2022;13:241.

45. Turner SL, Karahalios A, Forbes AB, Taljaard M, Grimshaw JM, McKenzie JE. Comparison of six statistical methods for interrupted time series studies: empirical evaluation of 190 published series. BMC Med Res Methodol. 2021;21:134.

46. Turner SL, Karahalios A, Forbes AB, Taljaard M, Grimshaw JM, Cheng AC, et al. Design characteristics and statistical methods used in interrupted time series studies evaluating public health interventions: a review. J Clin Epidemiol. 2020;122:1–11.

47. Chiarella C, He XZ, Hommes C. A dynamic analysis of moving average rules. J Econ Dyn Control. 2006;30:1729–53.

48. Thiyagarajan K, Kodagoda S, van Nguyen L. Predictive analytics for detecting sensor failure using autoregressive integrated moving average model. Proceedings of the 2017 12th IEEE Conference on Industrial Electronics and Applications, ICIEA 2017. 2018;2018-February:1926–31.

49. Akrami SA, El-Shafie A, Naseri M, Santos CAG. Rainfall data analyzing using moving average (MA) model and wavelet multi-resolution intelligent model for noise evaluation to improve the forecasting accuracy. Neural Computing and Applications 2014 25:7. 2014;25:1853–61.

